# Association of Egg Consumption, Metabolic Markers, and Risk of Cardiovascular Diseases

**DOI:** 10.1101/2021.12.06.21266787

**Authors:** Lang Pan, Lu Chen, Jun Lv, Yuanjie Pang, Yu Guo, Pei Pei, Huaidong Du, Ling Yang, Iona Y. Millwood, Robin G. Walters, Yiping Chen, Weiwei Gong, Junshi Chen, Canqing Yu, Zhengming Chen, Liming Li, the China Kadoorie Biobank Collaborative Group

## Abstract

**Objective:** To simultaneously explore the associations of self-reported egg consumption with plasma metabolic markers and these markers with CVD risk.

**Methods:** Totally 4,778 participants (3,401 CVD cases subdivided into subtypes and 1,377 controls) aged 30-79 were selected from a nested case-control study based on the China Kadoorie Biobank. Targeted nuclear magnetic resonance was used to quantify 225 metabolites and derived traits in baseline plasma samples. Linear regression was conducted to assess associations between self-reported egg consumption and metabolic markers, which were further compared with associations between metabolic markers and CVD risk.

**Results:** Egg consumption was associated with 24 out of 225 markers, including positive associations for apolipoprotein A1, acetate, mean HDL diameter, and lipid profiles of very large and large HDL, and inverse associations for total cholesterol and cholesterol esters in small VLDL. Among these 24 markers, 14 of them were associated with CVD risk. In general, the associations of metabolic markers with egg consumption and of metabolic markers with CVD risk showed opposite patterns.

**Conclusions:** In the Chinese population, egg consumption is associated with several metabolic markers, which may partially explain the protective effect of egg consumption on CVD.

## Introduction

Cardiovascular disease (CVD) has always been a major public health challenge and is the leading cause of death and disability worldwide, including in China.^[1]^ Ischemic heart disease (IHD), ischemic stroke (IS), and intracerebral hemorrhage (ICH) accounted for the majority of the deaths caused by CVD, which becomes more and more serious as the aggravation of population aging.^[1, 2]^ In recent decades, with the concern about the burden of CVD, researchers not only focus on the modifiable risk factors, including smoking, drinking, physical activity and diet, but also pay more and more attention to the internal biological mechanism.^[3, 4]^ Plasma lipids, especially low-density lipoprotein cholesterol (LDL-C) which may accumulate in the arterial wall, gradually form atherosclerotic plaques and block the corresponding artery, is generally considered to be associated with risks of IHD, IS and ICH.[5-7]

Eggs are one of the richest sources of dietary cholesterol. However, they also contain a wide variety of essential nutrients and bioactive compounds, such as high-quality protein, fat-soluble and B vitamins, phospholipids, and choline.^[8, 9]^ Evidence for the association between eggs and CVD remains controversial in both observational and prospective studies based on Western and Japanese populations, with positive or negative associations being reported in some studies ^[10]^ and others finding no significant association.^[11-16]^ Similar controversial findings have been found in the Chinese population. An inverse association between egg consumption and CVD risk was published,^[17]^ whereas more (>10 eggs per week) and less (<1 per week) egg intake was found to be harmful to cardiovascular health^[18]^. With regret, few studies have assessed the role of individual plasma cholesterol levels in such association, which may make the association clearer. Lipoproteins, on the other hand, can be divided into large, medium, and small subclasses according to particle size and include a series of constituents, including cholesterol, phospholipids, triglycerides, and apolipoprotein, which is difficult for conventional approaches to quantitate. Nuclear magnetic resonance (NMR) metabolomics provides a new opportunity to explore the associations between exposure, diseases, lipoprotein, and other small-molecule metabolic markers in a more detailed perspective.^[19]^

The present study aimed at simultaneously exploring the associations of self-reported egg consumption with plasma lipids, fatty acids, amino acids, and other metabolic markers, and of these markers with the risk of CVD in a nested case-control study in the China Kadoorie Biobank (CKB).

## Methods

### Participants and study design

The CKB study was a prospective cohort of 512,725 participants aged 30-79 years from 5 urban and 5 rural areas across China. Participants were recruited between 2004 and 2008, whose morbidity and mortality were followed up ever since. A laptop-based questionnaire was used to collect detailed information, including demographic characteristics (e.g., age at recruitment, sex, education, household income, and marital status), lifestyle factors (e.g., smoking and drinking habits, food intake, and physical activities), medical history (e.g., hypertension, diabetes, and use of certain specific medications such as statins), and family history of diabetes, heart attack or stroke. Each participant also underwent a range of physical measurements operated by trained staff, including anthropometry, lung function, blood pressure, and heart rate, etc. All participants provided a 10mL non-fasting (with time since last meal recorded) blood sample for immediate on-site random plasma glucose (RPG) test and long-term storage. In addition, every 5 years after completing the baseline survey, about 5% of the participants were randomly selected to join in the re-survey. Detailed descriptions of the CKB study have been previously published.^[20]^

The present study selected 4,778 participants from a previous nested case-control study based on CKB.^[21]^ Cases were consisted of incident cases of myocardial infarction (MI, ICD-10 I21-23, n=946), IS (I63 and I69.3, n=1,217), and ICH (I61 and I69.1, n=1,238), with a censoring date of 1 January 2015. And 1,377 controls were frequently matched to the combined cases by age, sex, and area if possible. All cases and controls had no history of self-reported prior doctor-diagnosed coronary heart disease (CHD), stroke, transient ischemic attack, or cancer and were not using statin therapy at baseline. The Ethical Review Committee of the Chinese Center for Disease Control and Prevention (Beijing, China) and the Oxford Tropical Research Ethics Committee, University of Oxford (UK) approved the study.

### Assessment of egg consumption

Using the brief food frequency questionnaire (FFQ) at baseline and the 1^st^ re-survey (2008), participants were asked about their frequency of habitual egg consumption as well as other 11 food groups (rice, wheat, other staple food, meat, poultry, fish, fresh vegetables, preserved vegetables, fresh fruits, soybean products, and dairy) during the past 12 months. Possible answers were “never/rarely, monthly, 1-3 days per week, 4-6 days per week, and daily”. The frequency was then converted into days of egg consumption per week, with each option corresponding to 0, 0.5, 2, 5, and 7 days per week, respectively. At the 2^nd^ re-survey (2013-2014), participants were additionally asked about the daily amount on days when consuming eggs.

### Measurement of NMR metabolisms

After centrifuging and aliquoting, baseline plasma samples from each participant were couriered from the regional laboratory via Beijing to Oxford for long-term storage in liquid nitrogen tanks. The stored plasma samples of the cases and controls were thawed and sub-aliquoted at the Wolfson laboratory, CTSU, before 100uL aliquots being shipped on dry ice to the Brainshake Laboratory at Oulu, Finland, for high-throughput targeted NMR spectroscopy to quantify 225 absolute concentrations or derived traits (e.g. lipids ratios) of metabolic markers simultaneously. Samples from cases and controls were quantified in a random order, with laboratory staff blinded to case or control status. The sample size of analyses of some metabolic markers involved was less than 4,778 since the quality control process rejected results of these metabolic markers among some participants.

### Statistical analysis

Baseline characteristics of participants were presented as means or percentages across controls and 3 subtypes of CVD cases, standardized by age, sex, and study region if appropriate, using multiple linear regressions for continuous variables or logistic regressions for categorical variables.

For metabolites whose measurements below the limit of detection were imputed with the lowest measured concentration. Each metabolic marker was log-transformed and divided by its standard deviation (SD). Linear regression was used to assess the associations of egg consumption with metabolic markers, adjusted for age, sex, region, education, household income, occupation, marital status, tea-drinking habit, smoking status, alcohol intake, physical activity, self-rated health, fasting time and frequency of other 11 food groups. For each biomarker, adjusted SD differences of log-transformed metabolic markers and 95% confidence intervals (CI) associated with an extra day of egg consumption per week were estimated.

In addition, participants were stratified by sex, age group (<60y or ≥60y), region (10 regions) and egg consumption group (never/rarely, monthly, 1-3 d/w, 4-6 d/w, or daily), and the mean values of amount on days when consuming eggs for each stratum collected in the 2^nd^ re-survey among 23,974 participants was calculated as a proxy to estimate the amount of egg consumption. The above calculation assumes that the daily amount of egg consumption for participants will not change significantly from baseline to the 2^nd^ re-survey.^[17]^

Logistic regression was used to estimate odds ratios (ORs) for CVD and its 3 subtypes (MI, IS, and ICH) per SD higher log-transformed metabolic markers, with the same variables adjusted for as in the analysis of egg consumption and metabolic markers. ICH cases were excluded from the analysis of metabolic markers and CVD because there were no associated metabolic markers with ICH.^[21]^ ORs were then plotted against SD differences in corresponding log-transformed metabolic biomarkers per extra day of egg consumption.

In order to examine the robustness of associations between egg consumption and metabolic markers, we performed several sensitivity analyses: additionally adjusting for body mass index (BMI), presence of prevalent hypertension or diabetes, and family history of diabetes or CVD; excluding participants with any metabolic markers below the limit of detection or rejected by quality control; using the weekly amount of egg consumption as a continuous independent variable instead of its frequency.

All *p*-values were two-sided, and statistical significance was defined as *p*<0.05. To account for a large number of highly correlated metabolic markers, we calculated the false discovery rate (FDR) *p*<0.05 based on the Benjamini-Hochberg method for associations of egg consumption with metabolic markers and of metabolic markers with risk of CVD. Statistical analyses were performed using Stata 15.0.

## Results

Age-, sex- and region-adjusted baseline characteristics of the 4,778 participants according to whether they developed CVD are shown in Table 1. Briefly, the mean (SD) age was 47.0 (8.2) years, 50.1% were women, and 29.0% resided in urban areas. The mean (SD) frequency of egg consumption and plasma total cholesterol concentration were 2.6 (2.3) days/week and 3.5 (0.6) mmol/L, respectively. Compared with controls, participants who subsequently developed any subtype of CVD were more likely to have poor self-rated health, to prevalent obesity, diabetes, and hypertension, and to have a family history of diabetes or CVD. Among them, MI or IS cases had a lower level of education but a higher level of household income, whereas ICH cases had higher levels of SBP and DBP.

**Table 1.** Baseline characteristics among 4,778 participants.

|  | Controls | MI or IS cases | ICH cases |
| --- | --- | --- | --- |
| N | 1,377 | 2,163 | 1,238 |
| Age, y | 46.83 (0.22) | 46.89 (0.18) | 47.34 (0.23) |
| Female, % | 50.29 (1.33) | 49.01 (1.07) | 51.72 (1.41) |
| Urban residents, % | 27.11 | 33.32 | 23.42 |
| Middle school or above, % | 56.97 | 53.76 | 56.36 |
| Income ≥35,000 RMB/year, % | 9.98 | 12.87 | 10.60 |
| Manual worker, % | 70.15 | 68.86 | 71.21 |
| BMI, kg/m<sup>2</sup> | 23.56 | 24.16 | 24.20 |
| Overweight/obesity, % | 43.45 (1.29) | 49.25 (1.04) | 49.98 (1.38) |
| SBP, mmHg | 127.94 | 136.68 | 149.76 |
| DBP, mmHg | 76.97 (0.36) | 82.34 (0.29) | 89.36 (0.39) |
| RBG, mmol/L | 5.63 (0.08) | 6.32 (0.06) | 6.31 (0.08) |
| Ever regular smoking, % | 34.63 (0.92) | 39.51 (0.68) | 36.04 (0.98) |
| Weekly drinking, % | 18.10 | 17.09 | 20.70 |
| Physical activity, MET h/d | 23.20 | 22.44 | 23.26 |
| Eggs consumption, d/w | 2.69 (0.06) | 2.54 (0.05) | 2.53 (0.06) |
| Fresh fruits consumption, d/w | 2.32 (0.05) | 2.00 (0.04) | 2.04 (0.06) |
| Red meat consumption, d/w | 3.26 (0.06) | 3.30 (0.04) | 3.31 (0.06) |
| ≥8h of fasting, % | 13.66 (0.86) | 15.98 (0.70) | 14.88 (0.99) |
| Poor self-rated health, % | 9.60 | 14.06 | 13.49 |
| Diabetes, % | 5.25 | 9.86 | 8.21 |
| Hypertension, % | 27.06 | 44.76 | 64.67 |
| Family history of diabetes, % | 6.83 | 8.74 | 7.71 |
| Family history of CVD, % | 23.54 | 26.96 | 30.98 |

Results were standardized by age, sex, and region where appropriate. Values are means
(standard errors, SE) or %.

Among the 225 metabolic markers or derived traits, 24 were associated with the frequency of egg consumption at FDR <5% (Figure 1). The associations for all 225 markers are shown in Table S1. Egg consumption was positively associated with lipoprotein particle concentrations of very large and large HDL. Similarly, within very large and large HDL, there were positive associations of total lipids, total cholesterol including its subclasses (cholesterol esters and free cholesterol), or phospholipids with egg consumption. Conversely, there was an inverse association of cholesterol esters in small VLDL with egg consumption. In addition to the absolute concentrations of lipids, the percentage of total cholesterol and cholesterol esters in large HDL were positively associated with egg consumption. Besides lipids, there were positive associations of acetate and apolipoprotein A1 with egg consumption, whereas an inverse association was observed for the ratio of apolipoprotein B to apolipoprotein A1.

**Figure 1.**
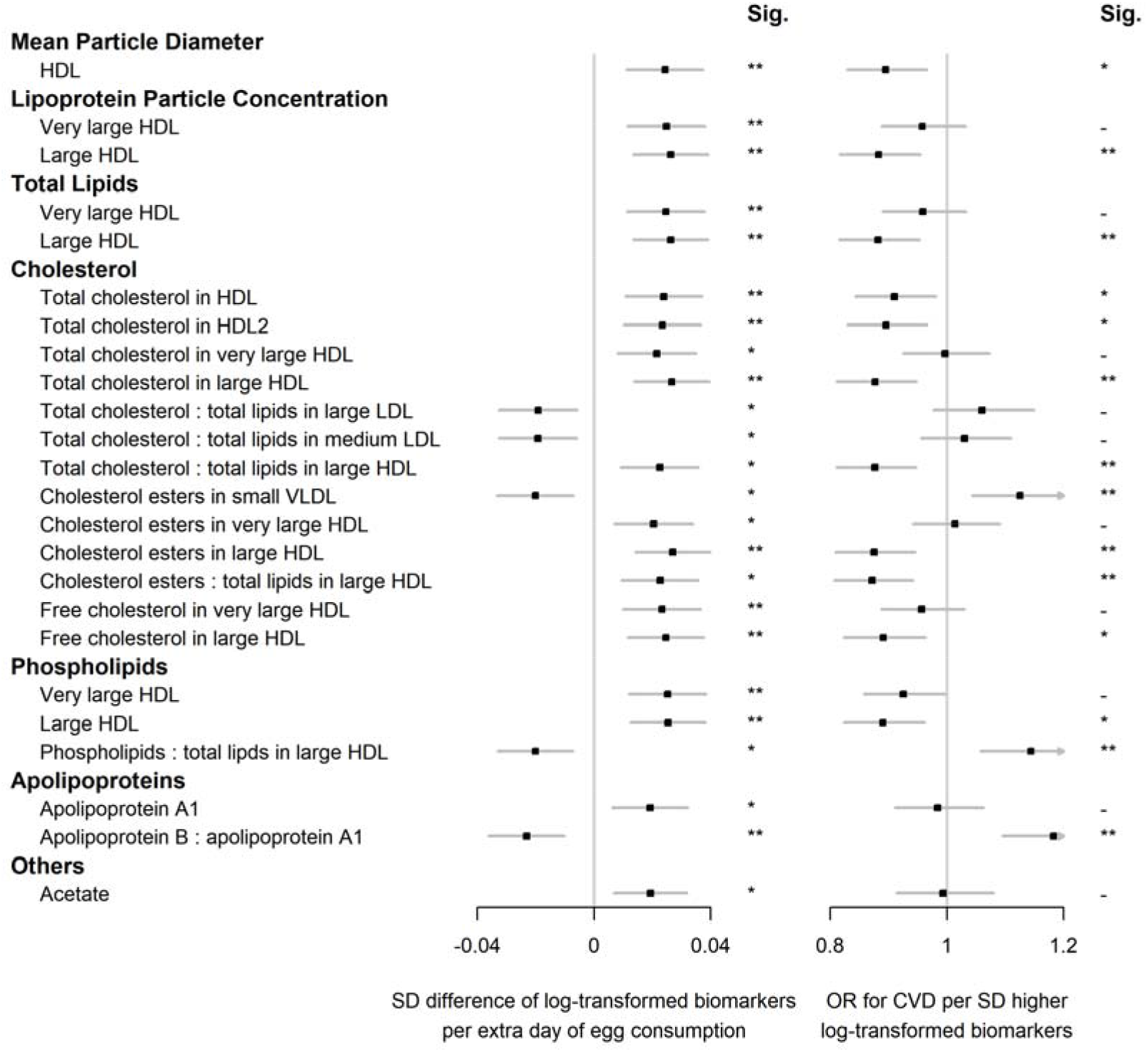
Significant associations of egg consumption and metabolic markers, and associations of these markers with risks of CVD. Models were adjusted for age, sex, region, education, household income, occupation, marital status, tea-drinking habit, smoking status, alcohol intake, physical activity, self-rated health, fasting time, and frequency of other 11 food groups. Black squares represented coefficients or ORs, while gray horizontal lines represented 95%CI. Significance (Sig.): \**p*<0.05, \*\**p*<0.01 (FDR-adjusted *p* using the Benjamini-Hochberg method).

Among 225 metabolic markers or derived traits, 96, 117, and 37 were associated with CVD and its subtypes, MI and IS, respectively, at FDR <5% (Table S1, Table S2). For total cholesterol within small VLDL, each SD increment in log-transformed value was associated with increased risks of CVD (OR 1.14 [95%CI 1.06-1.23]), MI (1.25 [1.13-1.38]), and IS (1.15 [1.04-1.26]), whereas the association with total cholesterol within large HDL was in the opposite direction (0.88 [0.81-0.95] for CVD and 0.83 [0.75-0.91] for MI). However, only one metabolic marker, glucose, showed a positive association with the risk of ICH. ORs for MI, IS, and ICH associated with all 225 metabolic markers were provided in Table S2. Among markers associated with risk of CVD, MI, or IS, 14, 15, and 2 were simultaneously associated with egg consumption. There was a clear pattern between the associations of egg consumption with metabolic markers and of these metabolic markers with disease risk; that is, metabolic markers associated with higher egg consumption tended to be associated with lower risk of CVD, MI, and IS (Pearson correlation: -0.72, -0.67, and -0.69, respectively; Table S1, Figure 2).

**Figure 2.**
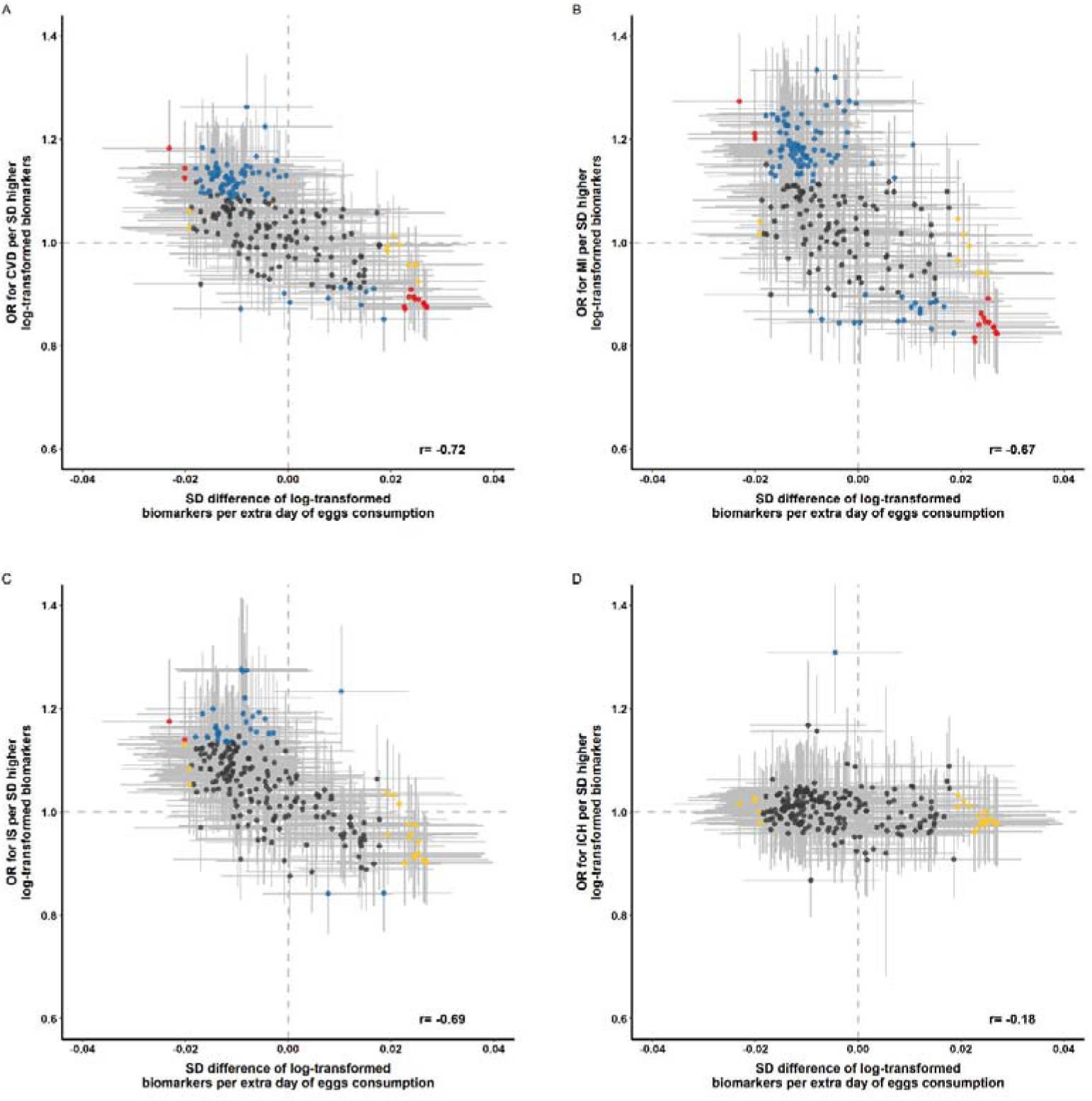
Global comparison of SD differences of 225 log-transformed metabolic markers associated with weekly days of egg consumption vs. ORs for (A) CVD, (B) MI, (C) IS, and (D) ICH associated with SD higher log-transformed metabolic markers. Models were adjusted for age, sex, region, education, household income, occupation, marital status, tea-drinking habit, smoking status, alcohol intake, physical activity, self-rated health, fasting time, and frequency of other 11 food groups. Yellow dots represented markers that were associated with egg consumption but not with risk of diseases; blue dots represented markers that were associated with risk of diseases but not with egg consumption; and red dots represented markers that were associated with both egg consumption and risk of diseases, with overlapping dots darker in color. The gray horizontal line and vertical line represented 95%CI of coefficients and ORs, respectively. Pearson correlations of coefficients and ORs were annotated in the lower right corner.

In sensitivity analyses, associations between egg consumption and metabolic markers remained essentially unchanged after further adjustment for hypertension, diabetes, and family history of diabetes or CVD (Figure S1). Similar associations were observed when restricting the analyses to participants without any metabolic markers below the detection limit or rejected by quality control (n=4,251) and using the weekly amount of egg consumption as a continuous independent variable instead of its frequency (Figure S2). However, when BMI was added to the basic model, the associations of egg consumption with metabolites were attenuated towards the null (Figure S1).

## Discussion

Based on data of FFQ and NMR metabolomics collected from the CKB population, we simultaneously assessed the associations between self-reported egg consumption and metabolic markers, and further explored whether these markers were associated with the risk of CVD and its subtypes. The results showed that egg consumption was positively associated with apolipoprotein A1, acetate, mean particle diameter of HDL, as well as lipid profiles in very large and large HDL including lipoprotein particle diameters, total lipids, total cholesterol, cholesterol esters, free cholesterol, and phospholipids. In contrast, inverse associations were observed in cholesterol esters in small VLDL, and the ratio of apolipoprotein B to apolipoprotein A1. Moreover, most of these metabolic markers were found to be inversely associated with the risk of CVD. On the whole, 225 metabolic markers showed a pattern; that is, their association with egg consumption and CVD risk were directionality opposite. Our results explained the protective effect of egg consumption on CVD risk through metabolism in the Chinese population.

Results from previous observational studies describing the association between egg consumption and the risk of CVD are inconsistent. However, so far, few studies have focused on plasma lipid metabolites between such associations. Some studies examined the metabonomics characteristics of plasma or feces in a wide range of food groups, including eggs, but not shown much interest in lipid metabolites.^[22, 23]^ Other studies combined eggs with other foods, such as ham, as a feed intervention to investigate consumers’ metabolomics characteristics, but the results can not reflect the metabolite changes caused by egg consumption alone.^[24, 25]^ This also gives expression to the necessity and urgency of the present study.

Even so, several studies and meta-analyses have explored the conventional lipids associated with egg consumption using blood biochemical analyses. Recently, a cohort study based on 39,021 overnight fasting adults in China collected data of egg consumption and blood lipids biochemical analysis.^[26]^ It was found that compared with the low egg consumption group (<26.79 g/d), medium or high consumption tertiles (>26.79 g/d) had higher levels of HDL-C, which was in line with the present study. It is worth noting that the mediating analysis of that study found that BMI or waist circumference (WC) could partly explain the association between egg consumption and HDL-C. The present study also supported the hypothesis according to sensitivity analysis, in which the associations were no longer significant by additional adjustment of BMI.

Consistent with our results, three small intervention studies based on Americans or Japanese observed positive effects for HDL-C and apolipoprotein A1.^[27-29]^ In addition, a meta-analysis of 66 eligible randomized clinical trials (RCT) involving 3,185 participants found a non-linear effect for VLDL-C, showing inverse association at the egg consumption level corresponding to the present study (<1.5 eggs/day).^[30]^

In our study, there was no significant association between egg consumption and lipoprotein in other sizes or densities except small VLDL, very large, and large HDL. To some extent, this reflected the balance between dietary cholesterol absorption and endogenous cholesterol biosynthesis, which accounts for about 25% and 75%, respectively,^[31]^ varying with different food consumption habits.^[31-33]^

Our study provides substantial evidence for the recommendation of egg consumption in current dietary guidelines in China. According to the Dietary Guidelines for Chinese Residents published in 2016, each standard adult should eat 40-50g eggs a day without discarding the yolk.^[34]^ However, according to the China Statistical Yearbook, the average egg purchase by Chinese residents in 2019 was 10.7 kilograms, or about 29.3 grams per day, which can be a partial estimate of actual consumption.^[35]^ Although this number has increased over the past decade, it still falls short of dietary guidelines. Hence, more health education and health promotion strategies and policies to encourage egg consumption need to be developed to improve lipid metabolite characteristics in the Chinese population, contributing to CVD prevention.

This study based on the CKB population has many strengths, including relatively large sample size, accurately identified CVD and its subtype events, collection of as many covariates as possible, and the quantification of a wide range of metabolites based on NMR platform, such as lipoproteins and their constituents with varies of size, density, and chemical structure. Our study also had several limitations. First, similar to other large-scale nutritional epidemiological studies, there was an unavoidable recall bias in the FFQ used for estimating egg and other food consumption in our study. There was also measurement bias when five options were used to estimate the weekly days of egg consumption. However, the results of our study were similar to those of previous observational studies or intervention studies, and these results remained robust when the imputed weekly amount of egg consumption was used as the independent variable. Second, even if potential confounding factors such as BMI, frequency of other food consumption, history of chronic diseases, and family history were adjusted in multivariate models or sensitivity analysis, residual confounding due to uncollected or suboptimally collected factors still existed. Third, as this study population’s weekly egg consumption (maximum 10.9 eggs per week) was not very large compared with other studies, it is necessary to be cautious when concluding this range. Finally, given the cross-sectional nature, the associations between baseline egg consumption and baseline level of plasma metabolites did not strongly elucidate their causality. Further longitudinal studies are needed to verify the causal roles of lipid metabolites in the association between egg consumption and CVD risk.

## Conclusions

This study set in the Chinese population found significant associations between egg consumption and acetate, lipid-related metabolites within very large and large HDL and small VLDL, and found that these associations were directionally opposite to associations between these metabolites and risk of CVD. These results we reported not only potentially reveal at the small molecule level that lipid metabolism metabolites may play a role in the beneficial effects of egg consumption on CVD but also provide Chinese population-based evidence for the formulation of strategies and policies to encourage egg consumption.

## Data Availability

All data produced are available online at www.ckbiobank.org/site/Data+Access.

## Acknowledgments

The most important acknowledgment is to the participants in the study and the members of the survey teams in each of the 10 regional centres, as well as to the project development and management teams based at Beijing, Oxford and the 10 regional centres.

## Funding

This work was supported by National Natural Science Foundation of China (81973125, 81941018, 91846303, 91843302). The CKB baseline survey and the first re-survey were supported by a grant from the Kadoorie Charitable Foundation in Hong Kong. The long-term follow-up is supported by grants (2016YFC0900500, 2016YFC0900501, 2016YFC0900504, 2016YFC1303904) from the National Key R&D Program of China, National Natural Science Foundation of China (81390540, 81390541, 81390544), and Chinese Ministry of Science and Technology (2011BAI09B01). The funders had no role in the study design, data collection, data analysis and interpretation, writing of the report, or the decision to submit the article for publication.

## Conflict of interest

none declared.

## Access to research materials/Data sharing

Details of how to access China Kadoorie Biobank data and details of the data release schedule are available from www.ckbiobank.org/site/Data+Access.

## Notes

### Competing Interest Statement

The authors have declared no competing interest.

### Funding Statement

This work was supported by the National Natural Science Foundation of China (81973125, 81941018, 91846303, 91843302). The CKB baseline survey and the first re-survey were supported by a grant from the Kadoorie Charitable Foundation in Hong Kong. The long-term follow-up is supported by grants (2016YFC0900500, 2016YFC0900501, 2016YFC0900504, 2016YFC1303904) from the National Key R&D Program of China, National Natural Science Foundation of China (81390540, 81390541, 81390544), and Chinese Ministry of Science and Technology (2011BAI09B01). The funders had no role in the study design, data collection, data analysis and interpretation, writing of the report, or the decision to submit the article for publication.

### Author Declarations

The Ethical Review Committee of the Chinese Center for Disease Control and Prevention (Beijing, China) and the Oxford Tropical Research Ethics Committee, University of Oxford (UK) approved the study.

