## Supplemental material and figures for "Association of Egg Consumption, Metabolic Markers, and Risk of Cardiovascular Diseases"

**International Steering Committee:** Junshi Chen, Zhengming Chen (PI), Robert Clarke, Rory Collins, Yu Guo, Liming Li (PI), Jun Lv, Richard Peto, Robin Walters. **International Co-ordinating Centre, Oxford:** Daniel Avery, Ruth Boxall, Derrick Bennett, Yumei Chang, Yiping Chen, Zhengming Chen, Robert Clarke, Huaidong Du, Simon Gilbert, Alex Hacker, Mike Hill, Michael Holmes, Andri Iona, Christiana Kartsonaki, Rene Kerosi, Ling Kong, Om Kurmi, Garry Lancaster, Sarah Lewington, Kuang Lin, John McDonnell, Iona Millwood, Qunhua Nie, Jayakrishnan Radhakrishnan, Paul Ryder, Sam Sansome, Dan Schmidt, Paul Sherliker, Rajani Sohoni, Becky Stevens, Iain Turnbull, Robin Walters, Jenny Wang, Lin Wang, Neil Wright, Ling Yang, Xiaoming Yang. **National Co-ordinating Centre, Beijing:** Yu Guo, Xiao Han, Can Hou, Jun Lv, Pei Pei, Chao Liu, Canqing Yu. **10 Regional Co-ordinating Centres: Qingdao CDC:** Zengchang Pang, Ruqin Gao, Shanpeng Li, Shaojie Wang, Yongmei Liu, Ranran Du, Yajing Zang, Liang Cheng, Xiaocao Tian, Hua Zhang, Yaoming Zhai, Feng Ning, Xiaohui Sun, Feifei Li. **Licang CDC:** Silu Lv, Junzheng Wang, Wei Hou. **Heilongjiang Provincial CDC:** Mingyuan Zeng, Ge Jiang, Xue Zhou. **Nangang CDC:** Liqiu Yang, Hui He, Bo Yu, Yanjie Li, Qinai Xu,Quan Kang, Ziyan Guo. **Hainan Provincial CDC:** Dan Wang, Ximin Hu, Jinyan Chen, Yan Fu, Zhenwang Fu, Xiaohuan Wang. **Meilan CDC:** Min Weng, Zhendong Guo, Shukuan Wu,Yilei Li, Huimei Li, Zhifang Fu. **Jiangsu Provincial CDC:** Ming Wu, Yonglin Zhou, Jinyi Zhou, Ran Tao, Jie Yang, Jian Su. **Suzhou CDC:** Fang liu, Jun Zhang, Yihe Hu, Yan Lu, , Liangcai Ma, Aiyu Tang, Shuo Zhang, Jianrong Jin, Jingchao Liu. **Guangxi Provincial CDC:** Zhenzhu Tang, Naying Chen, Ying Huang. **Liuzhou CDC:** Mingqiang Li, Jinhuai Meng, Rong Pan, Qilian Jiang, Jian Lan,Yun Liu, Liuping Wei, Liyuan Zhou, Ningyu Chen Ping Wang, Fanwen Meng, Yulu Qin,, Sisi Wang. **Sichuan Provincial CDC:** Xianping Wu, Ningmei Zhang, Xiaofang Chen,Weiwei Zhou. **Pengzhou CDC:** Guojin Luo, Jianguo Li, Xiaofang Chen, Xunfu Zhong, Jiaqiu Liu, Qiang Sun. **Gansu Provincial CDC:** Pengfei Ge, Xiaolan Ren, Caixia Dong. **Maiji CDC:** Hui Zhang, Enke Mao, Xiaoping Wang, Tao Wang, Xi zhang. **Henan Provincial CDC:** Ding Zhang, Gang Zhou, Shixian Feng, Liang Chang, Lei Fan. **Huixian CDC:** Yulian Gao, Tianyou He, Huarong Sun, Pan He, Chen Hu, Xukui Zhang, Huifang Wu, Pan He. **Zhejiang Provincial CDC:** Min Yu, Ruying Hu, Hao Wang. Tongxiang CDC: Yijian Qian, Chunmei Wang, Kaixu Xie, Lingli Chen, Yidan Zhang, Dongxia Pan, Qijun Gu. **Hunan Provincial CDC:** Yuelong Huang, Biyun Chen, Li Yin, Huilin Liu, Zhongxi Fu, Qiaohua Xu. **Liuyang CDC:** Xin Xu, Hao Zhang, Huajun Long, Xianzhi Li, Libo Zhang, Zhe Qiu.

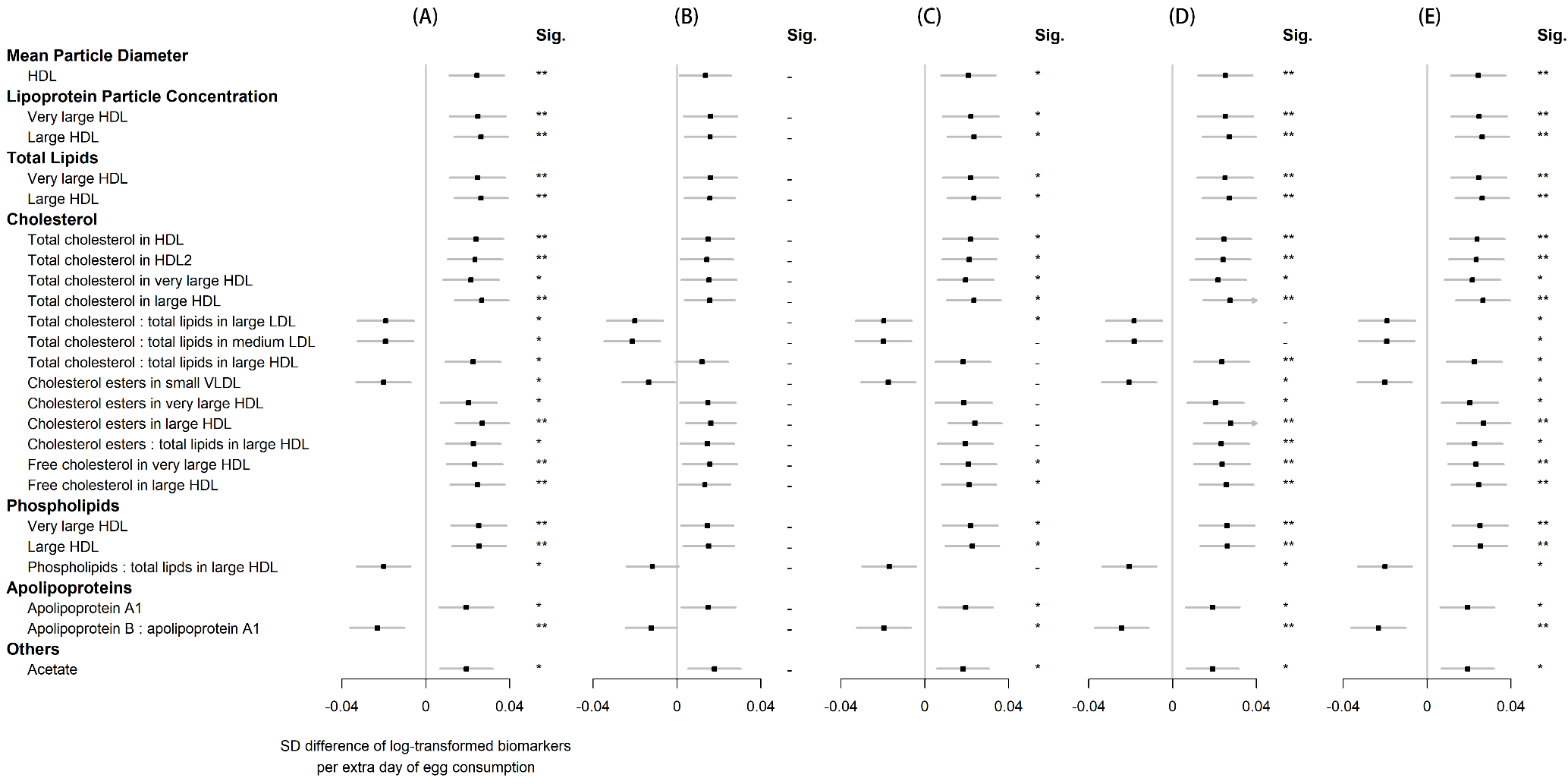

**Figure S1. Associations of egg consumption and log-transformed metabolic markers (A) in basic models, additionally adjusted for (B) BMI, (C) hypertension, (D) diabetes, and (E) family history of diabetes, heart attack or stroke. Significance (Sig.): * *p*<0.05, ** *p*<0.01 (FDR-adjusted *p* using the Benjamini-Hochberg method).**

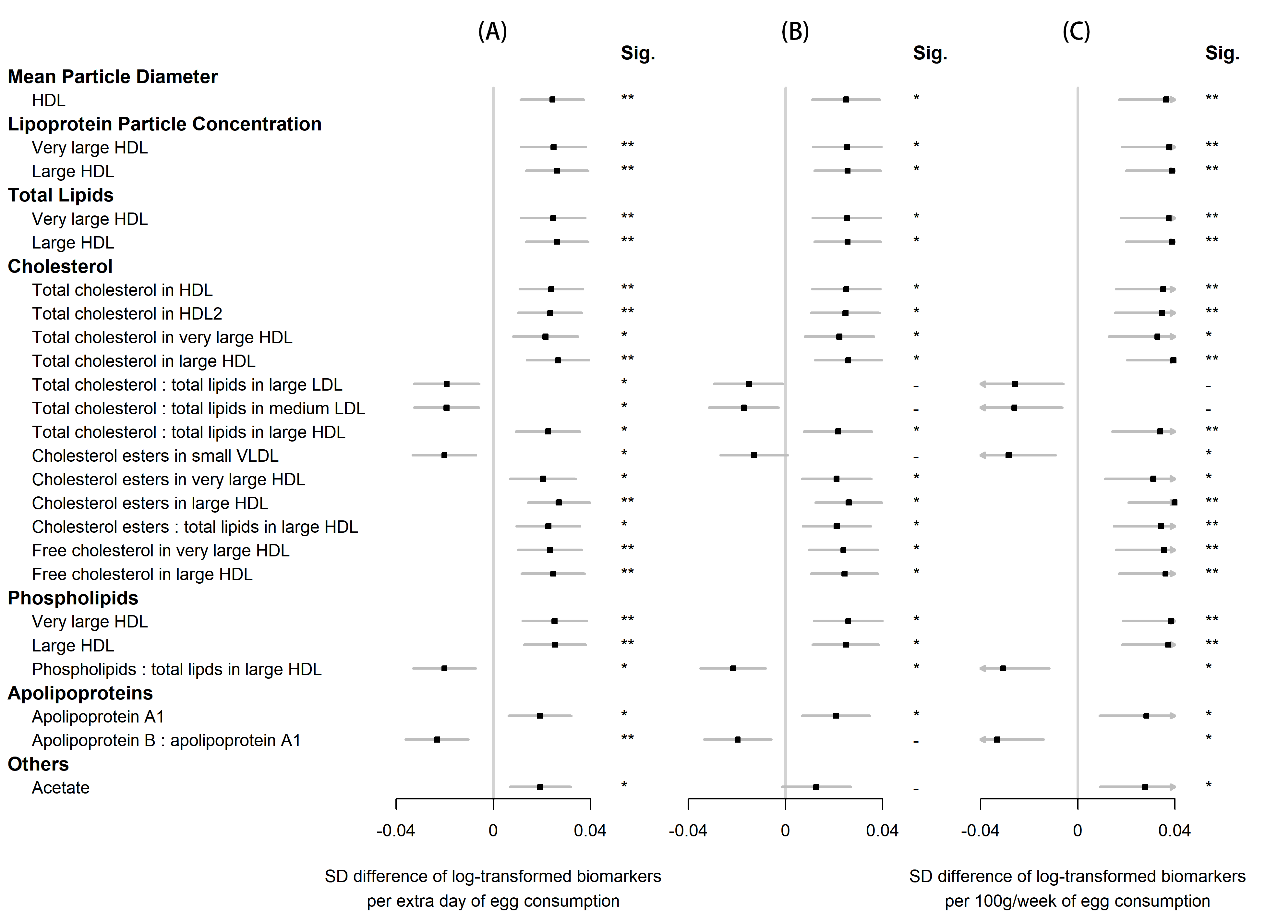

**Figure S2. Associations of egg consumption and log-transformed metabolic markers (A) in basic models, when (B) restricting the analyses to participants without any metabolic markers below the detection limit or rejected by the quality control (n=4,251), and (C) using weekly amount of egg consumption as continuous independent variable instead of its frequency.** **Significance (Sig.): * *p*<0.05, ** *p*<0.01 (FDR-adjusted *p* using the Benjamini-Hochberg method).**
