## Supplemental Table 1 for "Association of Egg Consumption, Metabolic Markers, and Risk of Cardiovascular Diseases"

**Table S1. Associations (95% CI) of egg consumption with all 225 log-transformed metabolic markers or devided traits, and of these markers with risks of CVD.**

| Metabolic marker | Unit | n | Coefficient (95% CI) | *p* | FDR-*p* | OR (95% CI) | *p* | FDR-*p* |
|---|---|---|---|---|---|---|---|---|
| Mean diameter for VLDL particles | nm | 4,777 | -0.005 (-0.018, 0.009) | 0.512 | 0.616 | 1.079 (1.003, 1.161) | 0.042 | 0.082 |
| Mean diameter for LDL particles | nm | 4,777 | 0.008 (-0.005, 0.020) | 0.228 | 0.333 | 0.892 (0.825, 0.965) | 0.004 | 0.015 |
| Mean diameter for HDL particles | nm | 4,777 | 0.024 (0.011, 0.037) | <0.001 | 0.006 | 0.895 (0.829, 0.965) | 0.004 | 0.015 |
| Concentration of chylomicrons and extremely large VLDL particles | mol/l | 4,778 | -0.011 (-0.024, 0.003) | 0.118 | 0.217 | 1.066 (0.993, 1.144) | 0.077 | 0.140 |
| Concentration of very large VLDL particles | mol/l | 4,778 | -0.008 (-0.021, 0.006) | 0.260 | 0.368 | 1.082 (1.006, 1.162) | 0.033 | 0.071 |
| Concentration of large VLDL particles | mol/l | 4,778 | -0.010 (-0.024, 0.003) | 0.139 | 0.243 | 1.109 (1.032, 1.192) | 0.005 | 0.017 |
| Concentration of medium VLDL particles | mol/l | 4,778 | -0.012 (-0.025, 0.002) | 0.082 | 0.193 | 1.126 (1.047, 1.212) | 0.001 | 0.008 |
| Concentration of small VLDL particles | mol/l | 4,778 | -0.016 (-0.029, -0.002) | 0.021 | 0.107 | 1.144 (1.063, 1.232) | <0.001 | 0.007 |
| Concentration of very small VLDL particles | mol/l | 4,778 | -0.014 (-0.027, -0.001) | 0.032 | 0.122 | 1.136 (1.054, 1.226) | 0.001 | 0.007 |
| Concentration of IDL particles | mol/l | 4,777 | -0.011 (-0.024, 0.002) | 0.102 | 0.205 | 1.099 (1.019, 1.186) | 0.014 | 0.040 |
| Concentration of large LDL particles | mol/l | 4,778 | -0.013 (-0.026, 0.000) | 0.056 | 0.159 | 1.112 (1.031, 1.200) | 0.006 | 0.019 |
| Concentration of medium LDL particles | mol/l | 4,778 | -0.015 (-0.028, -0.002) | 0.026 | 0.110 | 1.125 (1.044, 1.213) | 0.002 | 0.010 |
| Concentration of small LDL particles | mol/l | 4,778 | -0.013 (-0.027, 0.000) | 0.042 | 0.134 | 1.138 (1.055, 1.227) | 0.001 | 0.007 |
| Concentration of very large HDL particles | mol/l | 4,778 | 0.025 (0.012, 0.038) | <0.001 | 0.007 | 0.958 (0.889, 1.032) | 0.255 | 0.346 |
| Concentration of large HDL particles | mol/l | 4,778 | 0.026 (0.014, 0.039) | <0.001 | 0.004 | 0.883 (0.817, 0.954) | 0.002 | 0.008 |
| Concentration of medium HDL particles | mol/l | 4,778 | 0.012 (-0.001, 0.025) | 0.064 | 0.174 | 0.921 (0.852, 0.996) | 0.038 | 0.077 |
| Concentration of small HDL particles | mol/l | 4,778 | 0.002 (-0.011, 0.014) | 0.809 | 0.859 | 1.023 (0.947, 1.104) | 0.568 | 0.662 |
| Total lipids in chylomicrons and extremely large VLDL | mmol/l | 4,778 | -0.011 (-0.024, 0.003) | 0.118 | 0.216 | 1.067 (0.994, 1.145) | 0.073 | 0.135 |
| Total lipids in very large VLDL | mmol/l | 4,778 | -0.008 (-0.021, 0.006) | 0.259 | 0.369 | 1.081 (1.006, 1.162) | 0.033 | 0.070 |
| Total lipids in large VLDL | mmol/l | 4,778 | -0.010 (-0.024, 0.003) | 0.139 | 0.241 | 1.108 (1.030, 1.191) | 0.006 | 0.019 |
| Total lipids in medium VLDL | mmol/l | 4,778 | -0.012 (-0.025, 0.002) | 0.083 | 0.192 | 1.126 (1.046, 1.211) | 0.002 | 0.008 |
| Total lipids in small VLDL | mmol/l | 4,778 | -0.016 (-0.029, -0.002) | 0.020 | 0.106 | 1.146 (1.064, 1.234) | <0.001 | 0.008 |
| Total lipids in very small VLDL | mmol/l | 4,778 | -0.014 (-0.028, -0.001) | 0.030 | 0.118 | 1.129 (1.046, 1.217) | 0.002 | 0.008 |
| Total lipids in IDL | mmol/l | 4,777 | -0.011 (-0.024, 0.002) | 0.091 | 0.195 | 1.094 (1.014, 1.180) | 0.021 | 0.048 |
| Total lipids in large LDL | mmol/l | 4,778 | -0.013 (-0.026, 0.000) | 0.049 | 0.144 | 1.108 (1.027, 1.195) | 0.008 | 0.025 |
| Total lipids in medium LDL | mmol/l | 4,778 | -0.015 (-0.028, -0.002) | 0.025 | 0.109 | 1.122 (1.041, 1.210) | 0.003 | 0.011 |
| Total lipids in small LDL | mmol/l | 4,778 | -0.014 (-0.027, -0.001) | 0.040 | 0.131 | 1.132 (1.050, 1.221) | 0.001 | 0.008 |
| Total lipids in very large HDL | mmol/l | 4,778 | 0.025 (0.011, 0.038) | <0.001 | 0.006 | 0.958 (0.890, 1.033) | 0.264 | 0.356 |
| Total lipids in large HDL | mmol/l | 4,778 | 0.026 (0.014, 0.039) | <0.001 | 0.003 | 0.882 (0.816, 0.953) | 0.002 | 0.008 |
| Total lipids in medium HDL | mmol/l | 4,778 | 0.012 (-0.001, 0.025) | 0.066 | 0.172 | 0.919 (0.850, 0.993) | 0.033 | 0.069 |
| Total lipids in small HDL | mmol/l | 4,778 | 0.002 (-0.011, 0.014) | 0.818 | 0.864 | 1.019 (0.943, 1.100) | 0.637 | 0.731 |
| Serum total cholesterol | mmol/l | 4,777 | -0.005 (-0.018, 0.008) | 0.431 | 0.536 | 1.103 (1.023, 1.191) | 0.011 | 0.032 |
| Total cholesterol in VLDL | mmol/l | 4,777 | -0.014 (-0.027, 0.000) | 0.043 | 0.134 | 1.144 (1.062, 1.232) | <0.001 | 0.007 |
| Total cholesterol in chylomicrons and extremely large VLDL | mmol/l | 4,778 | -0.012 (-0.025, 0.002) | 0.086 | 0.194 | 1.068 (0.995, 1.147) | 0.067 | 0.125 |
| Total cholesterol in very large VLDL | mmol/l | 4,778 | -0.008 (-0.022, 0.005) | 0.218 | 0.323 | 1.064 (0.991, 1.143) | 0.088 | 0.154 |
| Total cholesterol in large VLDL | mmol/l | 4,778 | -0.011 (-0.024, 0.003) | 0.121 | 0.219 | 1.088 (1.013, 1.169) | 0.021 | 0.049 |

| | | | | | | | | |
|---|---|---|---|---|---|---|---|---|
| Total cholesterol in medium VLDL | mmol/l | 4,778 | -0.012 (-0.025, 0.002) | 0.086 | 0.195 | 1.114 (1.036, 1.198) | 0.003 | 0.014 |
| Total cholesterol in small VLDL | mmol/l | 4,778 | -0.018 (-0.031, -0.005) | 0.007 | 0.058 | 1.143 (1.061, 1.232) | <0.001 | 0.006 |
| Total cholesterol in very small VLDL | mmol/l | 4,778 | -0.015 (-0.028, -0.002) | 0.023 | 0.106 | 1.095 (1.015, 1.182) | 0.019 | 0.048 |
| Total cholesterol in IDL | mmol/l | 4,778 | -0.012 (-0.025, 0.002) | 0.083 | 0.192 | 1.084 (1.005, 1.169) | 0.038 | 0.077 |
| Total cholesterol in LDL | mmol/l | 4,777 | -0.017 (-0.030, -0.004) | 0.013 | 0.075 | 1.108 (1.025, 1.197) | 0.010 | 0.029 |
| Total cholesterol in large LDL | mmol/l | 4,778 | -0.015 (-0.028, -0.002) | 0.022 | 0.109 | 1.101 (1.021, 1.189) | 0.013 | 0.037 |
| Total cholesterol in medium LDL | mmol/l | 4,778 | -0.017 (-0.030, -0.004) | 0.011 | 0.070 | 1.097 (1.018, 1.183) | 0.015 | 0.040 |
| Total cholesterol in small LDL | mmol/l | 4,778 | -0.016 (-0.029, -0.003) | 0.016 | 0.085 | 1.096 (1.017, 1.181) | 0.016 | 0.042 |
| Total cholesterol in HDL | mmol/l | 4,777 | 0.024 (0.011, 0.037) | <0.001 | 0.007 | 0.910 (0.843, 0.981) | 0.014 | 0.040 |
| Total cholesterol in HDL2 | mmol/l | 4,777 | 0.023 (0.010, 0.037) | <0.001 | 0.008 | 0.895 (0.830, 0.966) | 0.004 | 0.015 |
| Total cholesterol in HDL3 | mmol/l | 4,777 | 0.017 (0.004, 0.030) | 0.010 | 0.068 | 1.058 (0.981, 1.141) | 0.147 | 0.218 |
| Total cholesterol in very large HDL | mmol/l | 4,778 | 0.022 (0.008, 0.035) | 0.002 | 0.021 | 0.996 (0.926, 1.073) | 0.925 | 0.938 |
| Total cholesterol in large HDL | mmol/l | 4,778 | 0.027 (0.014, 0.040) | <0.001 | 0.005 | 0.877 (0.811, 0.947) | 0.001 | 0.007 |
| Total cholesterol in medium HDL | mmol/l | 4,778 | 0.009 (-0.004, 0.022) | 0.183 | 0.293 | 0.914 (0.845, 0.988) | 0.023 | 0.053 |
| Total cholesterol in small HDL | mmol/l | 4,778 | -0.004 (-0.017, 0.009) | 0.525 | 0.625 | 1.034 (0.960, 1.115) | 0.375 | 0.476 |
| Remnant cholesterol (non-HDL, non-LDL -cholesterol) | mmol/l | 4,777 | -0.014 (-0.027, -0.001) | 0.033 | 0.121 | 1.146 (1.063, 1.237) | <0.001 | 0.006 |
| Esterified cholesterol | mmol/l | 4,774 | 0.005 (-0.008, 0.019) | 0.448 | 0.554 | 0.988 (0.903, 1.082) | 0.795 | 0.868 |
| Cholesterol esters in chylomicrons and extremely large VLDL | mmol/l | 4,778 | -0.013 (-0.026, 0.001) | 0.066 | 0.170 | 1.069 (0.995, 1.147) | 0.066 | 0.125 |
| Cholesterol esters in very large VLDL | mmol/l | 4,778 | -0.009 (-0.023, 0.004) | 0.188 | 0.295 | 1.056 (0.983, 1.134) | 0.134 | 0.204 |
| Cholesterol esters in large VLDL | mmol/l | 4,778 | -0.012 (-0.026, 0.002) | 0.086 | 0.192 | 1.062 (0.991, 1.139) | 0.090 | 0.156 |
| Cholesterol esters in medium VLDL | mmol/l | 4,778 | -0.013 (-0.026, 0.001) | 0.063 | 0.173 | 1.100 (1.022, 1.183) | 0.011 | 0.031 |
| Cholesterol esters in small VLDL | mmol/l | 4,778 | -0.020 (-0.033, -0.007) | 0.002 | 0.028 | 1.125 (1.044, 1.214) | 0.002 | 0.010 |
| Cholesterol esters in very small VLDL | mmol/l | 4,778 | -0.015 (-0.028, -0.002) | 0.023 | 0.104 | 1.090 (1.010, 1.177) | 0.027 | 0.060 |
| Cholesterol esters in IDL | mmol/l | 4,778 | -0.012 (-0.025, 0.001) | 0.081 | 0.194 | 1.097 (1.016, 1.184) | 0.017 | 0.044 |
| Cholesterol esters in large LDL | mmol/l | 4,778 | -0.017 (-0.030, -0.004) | 0.011 | 0.070 | 1.121 (1.035, 1.215) | 0.005 | 0.018 |
| Cholesterol esters in medium LDL | mmol/l | 4,778 | -0.019 (-0.032, -0.005) | 0.006 | 0.050 | 1.068 (0.992, 1.151) | 0.082 | 0.147 |
| Cholesterol esters in small LDL | mmol/l | 4,778 | -0.017 (-0.030, -0.004) | 0.013 | 0.075 | 1.060 (0.985, 1.142) | 0.122 | 0.193 |
| Cholesterol esters in very large HDL | mmol/l | 4,778 | 0.020 (0.007, 0.034) | 0.003 | 0.030 | 1.014 (0.942, 1.091) | 0.718 | 0.803 |
| Cholesterol esters in large HDL | mmol/l | 4,778 | 0.027 (0.014, 0.040) | <0.001 | 0.008 | 0.875 (0.809, 0.945) | 0.001 | 0.007 |
| Cholesterol esters in medium HDL | mmol/l | 4,778 | 0.008 (-0.005, 0.021) | 0.243 | 0.353 | 0.915 (0.846, 0.989) | 0.026 | 0.058 |
| Cholesterol esters in small HDL | mmol/l | 4,778 | -0.004 (-0.017, 0.010) | 0.593 | 0.677 | 1.031 (0.957, 1.111) | 0.426 | 0.535 |
| Free cholesterol | mmol/l | 4,774 | -0.014 (-0.027, -0.001) | 0.037 | 0.124 | 1.092 (1.013, 1.178) | 0.022 | 0.051 |
| Free cholesterol in chylomicrons and extremely large VLDL | mmol/l | 4,778 | -0.012 (-0.026, 0.002) | 0.081 | 0.193 | 1.064 (0.991, 1.142) | 0.087 | 0.154 |
| Free cholesterol in very large VLDL | mmol/l | 4,778 | -0.007 (-0.021, 0.006) | 0.305 | 0.409 | 1.058 (0.986, 1.136) | 0.119 | 0.190 |
| Free cholesterol in large VLDL | mmol/l | 4,778 | -0.011 (-0.025, 0.002) | 0.105 | 0.202 | 1.090 (1.015, 1.170) | 0.018 | 0.044 |
| Free cholesterol in medium VLDL | mmol/l | 4,778 | -0.012 (-0.025, 0.002) | 0.090 | 0.197 | 1.122 (1.043, 1.206) | 0.002 | 0.009 |
| Free cholesterol in small VLDL | mmol/l | 4,778 | -0.014 (-0.027, 0.000) | 0.043 | 0.132 | 1.143 (1.061, 1.231) | <0.001 | 0.007 |
| Free cholesterol in very small VLDL | mmol/l | 4,778 | -0.013 (-0.026, 0.000) | 0.059 | 0.165 | 1.097 (1.018, 1.182) | 0.015 | 0.041 |
| Free cholesterol in IDL | mmol/l | 4,778 | -0.011 (-0.025, 0.002) | 0.094 | 0.199 | 1.049 (0.973, 1.132) | 0.214 | 0.294 |

| | | | | | | | | |
|---|---|---|---|---|---|---|---|---|
| Free cholesterol in large LDL | mmol/l | 4,778 | -0.012 (-0.026, 0.001) | 0.068 | 0.171 | 1.070 (0.992, 1.154) | 0.081 | 0.146 |
| Free cholesterol in medium LDL | mmol/l | 4,778 | -0.011 (-0.024, 0.002) | 0.090 | 0.195 | 1.117 (1.036, 1.205) | 0.004 | 0.015 |
| Free cholesterol in small LDL | mmol/l | 4,778 | -0.011 (-0.024, 0.002) | 0.111 | 0.208 | 1.121 (1.040, 1.210) | 0.003 | 0.013 |
| Free cholesterol in very large HDL | mmol/l | 4,778 | 0.023 (0.010, 0.037) | 0.001 | 0.009 | 0.956 (0.888, 1.030) | 0.239 | 0.326 |
| Free cholesterol in large HDL | mmol/l | 4,778 | 0.025 (0.012, 0.038) | <0.001 | 0.006 | 0.891 (0.824, 0.963) | 0.004 | 0.014 |
| Free cholesterol in medium HDL | mmol/l | 4,778 | 0.012 (-0.001, 0.025) | 0.067 | 0.171 | 0.913 (0.845, 0.986) | 0.021 | 0.049 |
| Free cholesterol in small HDL | mmol/l | 4,778 | 0.008 (-0.005, 0.021) | 0.209 | 0.322 | 0.972 (0.899, 1.050) | 0.472 | 0.577 |
| Serum total triglycerides | mmol/l | 4,777 | -0.009 (-0.022, 0.005) | 0.213 | 0.318 | 1.131 (1.051, 1.217) | 0.001 | 0.007 |
| Triglycerides in VLDL | mmol/l | 4,777 | -0.011 (-0.024, 0.002) | 0.109 | 0.208 | 1.125 (1.046, 1.211) | 0.002 | 0.008 |
| Triglycerides in chylomicrons and extremely large VLDL | mmol/l | 4,778 | -0.011 (-0.025, 0.002) | 0.102 | 0.207 | 1.059 (0.987, 1.137) | 0.108 | 0.178 |
| Triglycerides in very large VLDL | mmol/l | 4,778 | -0.008 (-0.021, 0.006) | 0.266 | 0.375 | 1.081 (1.006, 1.162) | 0.034 | 0.070 |
| Triglycerides in large VLDL | mmol/l | 4,778 | -0.010 (-0.024, 0.003) | 0.142 | 0.242 | 1.113 (1.035, 1.196) | 0.004 | 0.015 |
| Triglycerides in medium VLDL | mmol/l | 4,778 | -0.012 (-0.025, 0.001) | 0.079 | 0.193 | 1.128 (1.048, 1.214) | 0.001 | 0.008 |
| Triglycerides in small VLDL | mmol/l | 4,778 | -0.015 (-0.029, -0.002) | 0.026 | 0.109 | 1.135 (1.054, 1.222) | 0.001 | 0.007 |
| Triglycerides in very small VLDL | mmol/l | 4,778 | -0.011 (-0.024, 0.002) | 0.104 | 0.205 | 1.135 (1.054, 1.222) | 0.001 | 0.007 |
| Triglycerides in IDL | mmol/l | 4,778 | -0.004 (-0.017, 0.009) | 0.560 | 0.653 | 1.132 (1.049, 1.222) | 0.001 | 0.008 |
| Triglycerides in LDL | mmol/l | 4,777 | 0.000 (-0.014, 0.013) | 0.950 | 0.967 | 1.131 (1.048, 1.220) | 0.002 | 0.008 |
| Triglycerides in large LDL | mmol/l | 4,778 | -0.002 (-0.015, 0.011) | 0.794 | 0.859 | 1.127 (1.044, 1.218) | 0.002 | 0.010 |
| Triglycerides in medium LDL | mmol/l | 4,778 | -0.003 (-0.016, 0.011) | 0.688 | 0.759 | 1.140 (1.058, 1.230) | 0.001 | 0.007 |
| Triglycerides in small LDL | mmol/l | 4,778 | -0.006 (-0.020, 0.007) | 0.363 | 0.480 | 1.167 (1.083, 1.257) | <0.001 | 0.002 |
| Triglycerides in HDL | mmol/l | 4,777 | 0.006 (-0.007, 0.019) | 0.372 | 0.484 | 1.050 (0.974, 1.133) | 0.201 | 0.279 |
| Triglycerides in very large HDL | mmol/l | 4,778 | 0.007 (-0.006, 0.020) | 0.287 | 0.394 | 1.063 (0.986, 1.147) | 0.112 | 0.182 |
| Triglycerides in large HDL | mmol/l | 4,778 | 0.014 (0.002, 0.027) | 0.024 | 0.105 | 0.973 (0.897, 1.054) | 0.499 | 0.597 |
| Triglycerides in medium HDL | mmol/l | 4,778 | -0.003 (-0.016, 0.011) | 0.700 | 0.768 | 1.038 (0.964, 1.117) | 0.325 | 0.425 |
| Triglycerides in small HDL | mmol/l | 4,778 | -0.004 (-0.018, 0.009) | 0.552 | 0.647 | 1.096 (1.018, 1.179) | 0.015 | 0.040 |
| Phospholipids in chylomicrons and extremely large VLDL | mmol/l | 4,778 | -0.012 (-0.025, 0.002) | 0.087 | 0.192 | 1.063 (0.991, 1.142) | 0.090 | 0.155 |
| Phospholipids in very large VLDL | mmol/l | 4,778 | -0.008 (-0.021, 0.006) | 0.277 | 0.385 | 1.059 (0.987, 1.137) | 0.111 | 0.181 |
| Phospholipids in large VLDL | mmol/l | 4,778 | -0.011 (-0.024, 0.003) | 0.125 | 0.222 | 1.107 (1.030, 1.190) | 0.006 | 0.018 |
| Phospholipids in medium VLDL | mmol/l | 4,778 | -0.012 (-0.026, 0.001) | 0.074 | 0.184 | 1.127 (1.047, 1.212) | 0.001 | 0.008 |
| Phospholipids in small VLDL | mmol/l | 4,778 | -0.014 (-0.027, -0.001) | 0.042 | 0.136 | 1.130 (1.050, 1.217) | 0.001 | 0.008 |
| Phospholipids in very small VLDL | mmol/l | 4,778 | -0.013 (-0.026, 0.000) | 0.052 | 0.150 | 1.104 (1.024, 1.191) | 0.010 | 0.030 |
| Phospholipids in IDL | mmol/l | 4,778 | -0.011 (-0.024, 0.002) | 0.096 | 0.199 | 1.077 (0.999, 1.162) | 0.053 | 0.101 |
| Phospholipids in large LDL | mmol/l | 4,778 | -0.012 (-0.025, 0.001) | 0.065 | 0.172 | 1.112 (1.030, 1.200) | 0.007 | 0.021 |
| Phospholipids in medium LDL | mmol/l | 4,778 | -0.012 (-0.025, 0.001) | 0.071 | 0.178 | 1.151 (1.066, 1.243) | <0.001 | 0.007 |
| Phospholipids in small LDL | mmol/l | 4,778 | -0.008 (-0.021, 0.005) | 0.213 | 0.319 | 1.149 (1.064, 1.240) | <0.001 | 0.007 |
| Phospholipids in very large HDL | mmol/l | 4,778 | 0.025 (0.012, 0.038) | <0.001 | 0.006 | 0.925 (0.858, 0.997) | 0.042 | 0.082 |
| Phospholipids in large HDL | mmol/l | 4,778 | 0.025 (0.013, 0.038) | <0.001 | 0.005 | 0.890 (0.824, 0.962) | 0.003 | 0.013 |
| Phospholipids in medium HDL | mmol/l | 4,778 | 0.015 (0.002, 0.027) | 0.022 | 0.108 | 0.924 (0.854, 0.999) | 0.047 | 0.089 |
| Phospholipids in small HDL | mmol/l | 4,778 | 0.006 (-0.007, 0.019) | 0.377 | 0.488 | 0.976 (0.905, 1.054) | 0.541 | 0.634 |

| | | | | | | | | |
|---|---|---|---|---|---|---|---|---|
| Apolipoprotein A-I | g/l | 4,777 | 0.019 (0.006, 0.032) | 0.003 | 0.035 | 0.984 (0.911, 1.062) | 0.678 | 0.766 |
| Apolipoprotein B | g/l | 4,775 | -0.015 (-0.028, -0.002) | 0.027 | 0.111 | 1.178 (1.091, 1.271) | <0.001 | 0.001 |
| Ratio of apolipoprotein B to apolipoprotein A-I | | 4,775 | -0.023 (-0.036, -0.010) | <0.001 | 0.008 | 1.183 (1.096, 1.277) | <0.001 | 0.001 |
| Total fatty acids | mmol/l | 4,769 | -0.008 (-0.022, 0.005) | 0.210 | 0.319 | 1.136 (1.053, 1.225) | 0.001 | 0.007 |
| Saturated fatty acids | mmol/l | 4,769 | -0.009 (-0.022, 0.004) | 0.194 | 0.304 | 1.117 (1.037, 1.203) | 0.004 | 0.014 |
| Monounsaturated fatty acids; 16:1, 18:1 | mmol/l | 4,769 | -0.007 (-0.020, 0.006) | 0.308 | 0.410 | 1.135 (1.053, 1.223) | 0.001 | 0.007 |
| Polyunsaturated fatty acids | mmol/l | 4,769 | -0.009 (-0.022, 0.004) | 0.178 | 0.288 | 1.141 (1.055, 1.233) | 0.001 | 0.007 |
| Omega-3 fatty acids | mmol/l | 4,769 | -0.006 (-0.018, 0.007) | 0.377 | 0.485 | 1.136 (1.050, 1.229) | 0.001 | 0.008 |
| 22:6, docosahexaenoic acid | mmol/l | 4,769 | -0.007 (-0.020, 0.005) | 0.259 | 0.371 | 1.147 (1.061, 1.239) | 0.001 | 0.007 |
| Omega-6 fatty acids | mmol/l | 4,769 | -0.009 (-0.022, 0.004) | 0.163 | 0.272 | 1.135 (1.051, 1.227) | 0.001 | 0.008 |
| 18:2, linoleic acid | mmol/l | 4,769 | -0.010 (-0.023, 0.004) | 0.165 | 0.273 | 0.979 (0.899, 1.066) | 0.626 | 0.722 |
| Ratio of saturated fatty acids to total fatty acids | % | 4,778 | 0.005 (-0.008, 0.018) | 0.487 | 0.596 | 0.919 (0.850, 0.993) | 0.033 | 0.071 |
| Ratio of monounsaturated fatty acids to total fatty acids | % | 4,778 | 0.003 (-0.010, 0.016) | 0.682 | 0.760 | 1.060 (0.984, 1.143) | 0.126 | 0.197 |
| Ratio of polyunsaturated fatty acids to total fatty acids | % | 4,778 | 0.000 (-0.013, 0.013) | 0.985 | 0.989 | 1.011 (0.938, 1.091) | 0.768 | 0.847 |
| Ratio of omega-3 fatty acids to total fatty acids | % | 4,778 | 0.003 (-0.009, 0.015) | 0.641 | 0.725 | 1.042 (0.963, 1.128) | 0.303 | 0.404 |
| Ratio of 22:6 docosahexaenoic acid to total fatty acids | % | 4,778 | 0.004 (-0.010, 0.017) | 0.592 | 0.679 | 1.009 (0.938, 1.084) | 0.816 | 0.870 |
| Ratio of omega-6 fatty acids to total fatty acids | % | 4,778 | 0.001 (-0.012, 0.014) | 0.928 | 0.957 | 1.005 (0.934, 1.082) | 0.895 | 0.920 |
| Ratio of 18:2 linoleic acid to total fatty acids | % | 4,778 | 0.000 (-0.014, 0.014) | 0.996 | 0.996 | 0.973 (0.902, 1.050) | 0.483 | 0.585 |
| Alanine | mmol/l | 4,776 | -0.010 (-0.024, 0.003) | 0.132 | 0.232 | 1.001 (0.931, 1.077) | 0.974 | 0.982 |
| Glutamine | mmol/l | 4,758 | -0.017 (-0.030, -0.004) | 0.011 | 0.068 | 0.919 (0.851, 0.993) | 0.033 | 0.070 |
| Histidine | mmol/l | 4,768 | -0.009 (-0.023, 0.004) | 0.184 | 0.292 | 0.994 (0.924, 1.069) | 0.866 | 0.907 |
| Isoleucine | mmol/l | 4,776 | 0.008 (-0.005, 0.021) | 0.209 | 0.320 | 1.009 (0.936, 1.088) | 0.815 | 0.873 |
| Leucine | mmol/l | 4,777 | 0.000 (-0.013, 0.012) | 0.954 | 0.967 | 1.007 (0.932, 1.089) | 0.855 | 0.903 |
| Phenylalanine | mmol/l | 4,777 | 0.018 (0.005, 0.030) | 0.007 | 0.057 | 0.993 (0.918, 1.075) | 0.869 | 0.905 |
| Tyrosine | mmol/l | 4,768 | 0.018 (0.004, 0.031) | 0.009 | 0.066 | 0.996 (0.923, 1.074) | 0.915 | 0.932 |
| Valine | mmol/l | 4,755 | 0.002 (-0.011, 0.015) | 0.798 | 0.859 | 1.000 (0.926, 1.080) | 0.998 | 0.998 |
| Citrate | mmol/l | 4,778 | 0.014 (0.000, 0.027) | 0.042 | 0.136 | 0.937 (0.869, 1.011) | 0.093 | 0.157 |
| Glucose | mmol/l | 4,766 | -0.005 (-0.018, 0.009) | 0.503 | 0.609 | 1.225 (1.132, 1.325) | <0.001 | <0.001 |
| Lactate | mmol/l | 4,777 | 0.001 (-0.012, 0.013) | 0.936 | 0.962 | 1.060 (0.982, 1.143) | 0.134 | 0.203 |
| Glycoprotein acetyls, mainly a1-acid glycoprotein | mmol/l | 4,777 | -0.008 (-0.021, 0.005) | 0.222 | 0.326 | 1.263 (1.167, 1.366) | <0.001 | <0.001 |
| Acetoacetate | mmol/l | 4,777 | 0.010 (-0.003, 0.023) | 0.122 | 0.220 | 1.050 (0.974, 1.132) | 0.206 | 0.284 |
| Acetate | mmol/l | 4,777 | 0.019 (0.007, 0.032) | 0.002 | 0.029 | 0.993 (0.914, 1.080) | 0.878 | 0.910 |
| 3-hydroxybutyrate | mmol/l | 4,667 | -0.002 (-0.015, 0.010) | 0.736 | 0.800 | 1.159 (1.065, 1.261) | 0.001 | 0.007 |
| Cholesterol esters to total lipids ratio in IDL | % | 4,778 | -0.011 (-0.024, 0.002) | 0.109 | 0.206 | 1.066 (0.987, 1.150) | 0.102 | 0.170 |
| Cholesterol esters to total lipids ratio in chylomicrons and extremely large VLDL | % | 4,778 | -0.015 (-0.028, -0.001) | 0.037 | 0.124 | 1.054 (0.982, 1.132) | 0.143 | 0.215 |
| Cholesterol esters to total lipids ratio in large HDL | % | 4,778 | 0.023 (0.010, 0.036) | 0.001 | 0.011 | 0.872 (0.807, 0.942) | <0.001 | 0.007 |
| Cholesterol esters to total lipids ratio in large LDL | % | 4,778 | -0.018 (-0.031, -0.004) | 0.010 | 0.068 | 1.117 (1.011, 1.234) | 0.030 | 0.065 |
| Cholesterol esters to total lipids ratio in large VLDL | % | 4,778 | -0.009 (-0.023, 0.004) | 0.183 | 0.292 | 0.999 (0.933, 1.070) | 0.976 | 0.981 |
| Cholesterol esters to total lipids ratio in medium HDL | % | 4,778 | -0.007 (-0.020, 0.006) | 0.291 | 0.397 | 0.945 (0.871, 1.025) | 0.171 | 0.249 |

| | | | | | | | | |
|---|---|---|---|---|---|---|---|---|
| Cholesterol esters to total lipids ratio in medium LDL | % | 4,778 | -0.018 (-0.032, -0.005) | 0.007 | 0.057 | 1.028 (0.954, 1.108) | 0.467 | 0.574 |
| Cholesterol esters to total lipids ratio in medium VLDL | % | 4,778 | -0.003 (-0.016, 0.010) | 0.624 | 0.709 | 0.950 (0.879, 1.027) | 0.200 | 0.281 |
| Cholesterol esters to total lipids ratio in small HDL | % | 4,778 | -0.004 (-0.018, 0.009) | 0.548 | 0.645 | 1.028 (0.953, 1.108) | 0.480 | 0.584 |
| Cholesterol esters to total lipids ratio in small LDL | % | 4,778 | -0.016 (-0.029, -0.002) | 0.021 | 0.106 | 1.022 (0.948, 1.101) | 0.572 | 0.664 |
| Cholesterol esters to total lipids ratio in small VLDL | % | 4,778 | -0.011 (-0.024, 0.003) | 0.114 | 0.212 | 0.994 (0.922, 1.073) | 0.883 | 0.911 |
| Cholesterol esters to total lipids ratio in very large HDL | % | 4,778 | -0.017 (-0.029, -0.004) | 0.010 | 0.067 | 1.184 (1.095, 1.280) | <0.001 | 0.001 |
| Cholesterol esters to total lipids ratio in very large VLDL | % | 4,778 | -0.008 (-0.021, 0.006) | 0.280 | 0.387 | 0.991 (0.923, 1.064) | 0.808 | 0.874 |
| Cholesterol esters to total lipids ratio in very small VLDL | % | 4,778 | -0.005 (-0.018, 0.008) | 0.478 | 0.587 | 0.936 (0.866, 1.011) | 0.093 | 0.156 |
| Free cholesterol to total lipids ratio in IDL | % | 4,778 | -0.008 (-0.021, 0.006) | 0.257 | 0.370 | 0.969 (0.892, 1.053) | 0.456 | 0.564 |
| Free cholesterol to total lipids ratio in chylomicrons and extremely large VLDL | % | 4,778 | -0.015 (-0.028, -0.001) | 0.035 | 0.123 | 1.049 (0.978, 1.126) | 0.180 | 0.259 |
| Free cholesterol to total lipids ratio in large HDL | % | 4,778 | 0.015 (0.001, 0.028) | 0.033 | 0.121 | 0.937 (0.863, 1.018) | 0.122 | 0.192 |
| Free cholesterol to total lipids ratio in large LDL | % | 4,778 | 0.000 (-0.013, 0.014) | 0.963 | 0.972 | 0.884 (0.815, 0.960) | 0.003 | 0.013 |
| Free cholesterol to total lipids ratio in large VLDL | % | 4,778 | -0.011 (-0.025, 0.002) | 0.101 | 0.206 | 1.055 (0.984, 1.131) | 0.133 | 0.204 |
| Free cholesterol to total lipids ratio in medium HDL | % | 4,778 | 0.010 (-0.003, 0.023) | 0.127 | 0.226 | 0.913 (0.845, 0.986) | 0.020 | 0.049 |
| Free cholesterol to total lipids ratio in medium LDL | % | 4,778 | 0.017 (0.004, 0.030) | 0.013 | 0.073 | 0.911 (0.846, 0.982) | 0.015 | 0.040 |
| Free cholesterol to total lipids ratio in medium VLDL | % | 4,778 | -0.009 (-0.022, 0.005) | 0.211 | 0.319 | 1.091 (1.016, 1.173) | 0.017 | 0.044 |
| Free cholesterol to total lipids ratio in small HDL | % | 4,778 | 0.014 (0.001, 0.027) | 0.031 | 0.121 | 0.914 (0.845, 0.988) | 0.024 | 0.055 |
| Free cholesterol to total lipids ratio in small LDL | % | 4,778 | 0.015 (0.002, 0.028) | 0.023 | 0.106 | 0.905 (0.839, 0.976) | 0.010 | 0.030 |
| Free cholesterol to total lipids ratio in small VLDL | % | 4,778 | 0.014 (0.001, 0.027) | 0.035 | 0.122 | 0.974 (0.903, 1.051) | 0.499 | 0.600 |
| Free cholesterol to total lipids ratio in very large HDL | % | 4,778 | 0.006 (-0.008, 0.019) | 0.416 | 0.523 | 0.966 (0.898, 1.039) | 0.353 | 0.454 |
| Free cholesterol to total lipids ratio in very large VLDL | % | 4,778 | -0.006 (-0.019, 0.008) | 0.402 | 0.508 | 1.034 (0.963, 1.109) | 0.356 | 0.456 |
| Free cholesterol to total lipids ratio in very small VLDL | % | 4,778 | -0.003 (-0.016, 0.011) | 0.688 | 0.763 | 0.996 (0.919, 1.078) | 0.913 | 0.933 |
| Phospholipids to total lipds ratio in IDL | % | 4,778 | -0.001 (-0.014, 0.013) | 0.912 | 0.950 | 0.901 (0.832, 0.976) | 0.010 | 0.030 |
| Phospholipids to total lipds ratio in chylomicrons and extremely large VLDL | % | 4,778 | -0.014 (-0.028, 0.000) | 0.045 | 0.134 | 1.048 (0.977, 1.124) | 0.193 | 0.274 |
| Phospholipids to total lipds ratio in large HDL | % | 4,778 | -0.020 (-0.033, -0.007) | 0.002 | 0.027 | 1.144 (1.058, 1.237) | 0.001 | 0.007 |
| Phospholipids to total lipds ratio in large LDL | % | 4,778 | 0.011 (-0.002, 0.024) | 0.105 | 0.204 | 0.928 (0.863, 0.998) | 0.044 | 0.086 |
| Phospholipids to total lipds ratio in large VLDL | % | 4,778 | -0.012 (-0.026, 0.001) | 0.081 | 0.195 | 1.081 (1.008, 1.160) | 0.030 | 0.066 |
| Phospholipids to total lipds ratio in medium HDL | % | 4,778 | 0.011 (-0.002, 0.023) | 0.103 | 0.206 | 1.065 (0.981, 1.155) | 0.132 | 0.204 |
| Phospholipids to total lipds ratio in medium LDL | % | 4,778 | 0.013 (-0.001, 0.026) | 0.060 | 0.166 | 0.963 (0.896, 1.036) | 0.311 | 0.410 |
| Phospholipids to total lipds ratio in medium VLDL | % | 4,778 | -0.005 (-0.018, 0.009) | 0.502 | 0.611 | 0.976 (0.907, 1.050) | 0.517 | 0.616 |
| Phospholipids to total lipds ratio in small HDL | % | 4,778 | 0.008 (-0.005, 0.022) | 0.207 | 0.321 | 0.937 (0.870, 1.010) | 0.091 | 0.155 |
| Phospholipids to total lipds ratio in small LDL | % | 4,778 | 0.015 (0.001, 0.028) | 0.029 | 0.118 | 0.947 (0.881, 1.019) | 0.148 | 0.219 |
| Phospholipids to total lipds ratio in small VLDL | % | 4,778 | 0.019 (0.005, 0.032) | 0.006 | 0.051 | 0.851 (0.790, 0.917) | <0.001 | 0.001 |
| Phospholipids to total lipds ratio in very large HDL | % | 4,778 | 0.014 (0.001, 0.027) | 0.033 | 0.119 | 0.879 (0.813, 0.952) | 0.001 | 0.008 |
| Phospholipids to total lipds ratio in very large VLDL | % | 4,778 | -0.006 (-0.020, 0.007) | 0.372 | 0.487 | 1.022 (0.953, 1.096) | 0.540 | 0.636 |
| Phospholipids to total lipds ratio in very small VLDL | % | 4,778 | -0.004 (-0.018, 0.009) | 0.534 | 0.632 | 1.012 (0.936, 1.093) | 0.769 | 0.844 |
| Total cholesterol to total lipids ratio in IDL | % | 4,778 | -0.014 (-0.027, 0.000) | 0.044 | 0.133 | 1.017 (0.940, 1.100) | 0.672 | 0.764 |
| Total cholesterol to total lipids ratio in chylomicrons and extremely large VLDL | % | 4,778 | -0.014 (-0.027, 0.000) | 0.050 | 0.147 | 1.056 (0.984, 1.134) | 0.130 | 0.202 |
| Total cholesterol to total lipids ratio in large HDL | % | 4,778 | 0.023 (0.009, 0.036) | 0.001 | 0.012 | 0.876 (0.811, 0.947) | 0.001 | 0.007 |

| | | | | | | | | |
|---|---|---|---|---|---|---|---|---|
| Total cholesterol to total lipids ratio in large LDL | % | 4,778 | -0.019 (-0.033, -0.006) | 0.005 | 0.046 | 1.060 (0.977, 1.150) | 0.160 | 0.233 |
| Total cholesterol to total lipids ratio in large VLDL | % | 4,778 | -0.010 (-0.024, 0.004) | 0.148 | 0.250 | 1.014 (0.946, 1.087) | 0.687 | 0.773 |
| Total cholesterol to total lipids ratio in medium HDL | % | 4,778 | -0.004 (-0.017, 0.010) | 0.584 | 0.673 | 0.930 (0.858, 1.007) | 0.074 | 0.136 |
| Total cholesterol to total lipids ratio in medium LDL | % | 4,778 | -0.019 (-0.033, -0.006) | 0.005 | 0.045 | 1.030 (0.956, 1.110) | 0.435 | 0.543 |
| Total cholesterol to total lipids ratio in medium VLDL | % | 4,778 | -0.003 (-0.016, 0.010) | 0.660 | 0.743 | 0.991 (0.919, 1.069) | 0.824 | 0.874 |
| Total cholesterol to total lipids ratio in small HDL | % | 4,778 | -0.006 (-0.019, 0.008) | 0.395 | 0.503 | 1.029 (0.955, 1.109) | 0.448 | 0.556 |
| Total cholesterol to total lipids ratio in small LDL | % | 4,778 | -0.018 (-0.031, -0.004) | 0.009 | 0.064 | 1.024 (0.950, 1.102) | 0.538 | 0.637 |
| Total cholesterol to total lipids ratio in small VLDL | % | 4,778 | -0.006 (-0.019, 0.007) | 0.363 | 0.478 | 0.990 (0.919, 1.067) | 0.800 | 0.870 |
| Total cholesterol to total lipids ratio in very large HDL | % | 4,778 | -0.014 (-0.027, -0.001) | 0.032 | 0.121 | 1.153 (1.067, 1.245) | <0.001 | 0.008 |
| Total cholesterol to total lipids ratio in very large VLDL | % | 4,778 | -0.007 (-0.021, 0.006) | 0.298 | 0.404 | 1.009 (0.940, 1.083) | 0.811 | 0.873 |
| Total cholesterol to total lipids ratio in very small VLDL | % | 4,778 | -0.006 (-0.019, 0.007) | 0.384 | 0.491 | 0.937 (0.865, 1.015) | 0.112 | 0.181 |
| Triglycerides to total lipids ratio in IDL | % | 4,778 | 0.007 (-0.006, 0.020) | 0.300 | 0.404 | 1.038 (0.962, 1.120) | 0.336 | 0.434 |
| Triglycerides to total lipids ratio in chylomicrons and extremely large VLDL | % | 4,778 | -0.011 (-0.025, 0.002) | 0.104 | 0.204 | 1.036 (0.966, 1.111) | 0.325 | 0.423 |
| Triglycerides to total lipids ratio in large HDL | % | 4,778 | -0.003 (-0.015, 0.010) | 0.703 | 0.768 | 1.068 (0.990, 1.152) | 0.091 | 0.156 |
| Triglycerides to total lipids ratio in large LDL | % | 4,778 | 0.011 (-0.002, 0.024) | 0.096 | 0.200 | 1.012 (0.937, 1.094) | 0.753 | 0.835 |
| Triglycerides to total lipids ratio in large VLDL | % | 4,778 | -0.001 (-0.014, 0.013) | 0.926 | 0.960 | 0.993 (0.924, 1.068) | 0.856 | 0.900 |
| Triglycerides to total lipids ratio in medium HDL | % | 4,778 | -0.009 (-0.023, 0.004) | 0.175 | 0.285 | 1.081 (1.005, 1.163) | 0.037 | 0.075 |
| Triglycerides to total lipids ratio in medium LDL | % | 4,778 | 0.012 (-0.001, 0.025) | 0.064 | 0.173 | 1.017 (0.943, 1.098) | 0.662 | 0.756 |
| Triglycerides to total lipids ratio in medium VLDL | % | 4,778 | -0.002 (-0.015, 0.011) | 0.802 | 0.860 | 1.013 (0.939, 1.094) | 0.735 | 0.819 |
| Triglycerides to total lipids ratio in small HDL | % | 4,778 | -0.004 (-0.018, 0.009) | 0.513 | 0.614 | 1.092 (1.014, 1.175) | 0.019 | 0.048 |
| Triglycerides to total lipids ratio in small LDL | % | 4,778 | 0.006 (-0.008, 0.019) | 0.418 | 0.522 | 1.054 (0.979, 1.135) | 0.159 | 0.235 |
| Triglycerides to total lipids ratio in small VLDL | % | 4,778 | -0.007 (-0.021, 0.006) | 0.267 | 0.373 | 1.050 (0.974, 1.131) | 0.200 | 0.280 |
| Triglycerides to total lipids ratio in very large HDL | % | 4,778 | -0.010 (-0.023, 0.003) | 0.141 | 0.242 | 1.094 (1.017, 1.177) | 0.016 | 0.042 |
| Triglycerides to total lipids ratio in very large VLDL | % | 4,778 | -0.002 (-0.015, 0.012) | 0.806 | 0.859 | 0.954 (0.888, 1.025) | 0.195 | 0.276 |
| Triglycerides to total lipids ratio in very small VLDL | % | 4,778 | 0.001 (-0.012, 0.015) | 0.848 | 0.887 | 1.039 (0.965, 1.120) | 0.310 | 0.410 |
| Total cholines | mmol/l | 4,771 | -0.004 (-0.017, 0.010) | 0.580 | 0.673 | 1.076 (0.999, 1.159) | 0.054 | 0.102 |
| Phosphatidylcholine and other cholines | mmol/l | 4,771 | -0.001 (-0.014, 0.013) | 0.940 | 0.962 | 1.050 (0.976, 1.131) | 0.191 | 0.274 |
| Total phosphoglycerides | mmol/l | 4,771 | -0.003 (-0.016, 0.010) | 0.669 | 0.749 | 1.082 (1.004, 1.166) | 0.038 | 0.076 |
| Ratio of triglycerides to phosphoglycerides | | 4,777 | -0.010 (-0.023, 0.004) | 0.154 | 0.259 | 1.114 (1.036, 1.199) | 0.004 | 0.014 |
| Albumin | signal area | 4,778 | -0.009 (-0.022, 0.004) | 0.167 | 0.274 | 0.872 (0.806, 0.943) | 0.001 | 0.007 |
| Creatinine | mmol/l | 4,746 | -0.010 (-0.021, 0.002) | 0.091 | 0.194 | 1.087 (0.983, 1.201) | 0.105 | 0.173 |
| Estimated degree of unsaturation | | 4,778 | 0.001 (-0.012, 0.014) | 0.836 | 0.879 | 0.969 (0.898, 1.045) | 0.417 | 0.527 |
| Sphingomyelins | mmol/l | 4,771 | -0.016 (-0.029, -0.003) | 0.014 | 0.077 | 1.042 (0.967, 1.123) | 0.276 | 0.370 |

Models were adjusted for age, sex, region, education, household income, occupation, marital status, tea drinking habit, smoking status, alcohol intake, physical activity, self-rated health, fasting time and frequency of other 11 food groups. Sample size of analyses (n) of some metabolic markers involved was less than 4,778, since results of these metabolic markers were rejected by the quality control process among some participants.
