## Supplemental Table 2 for "Association of Egg Consumption, Metabolic Markers, and Risk of Cardiovascular Diseases"

**Table S2. ORs (95% CI) for MI, IS, and ICH per SD higher log-transformed metabolic markers.**

| Metabolic marker | Unit | OR (95% CI) for MI | *p* | FDR-*p* | OR (95% CI) for IS | *p* | FDR-*p* | OR (95% CI) for ICH | *p* | FDR-*p* |
|---|---|---|---|---|---|---|---|---|---|---|
| Mean diameter for VLDL particles | nm | 1.091 (0.991, 1.200) | 0.076 | 0.121 | 1.102 (1.008, 1.205) | 0.033 | 0.098 | 1.036 (0.952, 1.126) | 0.412 | 1.570 |
| Mean diameter for LDL particles | nm | 0.904 (0.817, 1.001) | 0.053 | 0.091 | 0.842 (0.763, 0.930) | 0.001 | 0.013 | 0.958 (0.879, 1.045) | 0.333 | 2.269 |
| Mean diameter for HDL particles | nm | 0.854 (0.774, 0.943) | 0.002 | 0.005 | 0.913 (0.831, 1.003) | 0.058 | 0.145 | 0.983 (0.904, 1.069) | 0.685 | 1.064 |
| Concentration of chylomicrons and extremely large VLDL particles | mol/l | 1.097 (0.993, 1.211) | 0.068 | 0.109 | 1.078 (0.990, 1.175) | 0.084 | 0.181 | 1.014 (0.935, 1.100) | 0.743 | 1.045 |
| Concentration of very large VLDL particles | mol/l | 1.113 (1.011, 1.226) | 0.029 | 0.055 | 1.104 (1.010, 1.207) | 0.030 | 0.094 | 1.023 (0.941, 1.113) | 0.592 | 1.280 |
| Concentration of large VLDL particles | mol/l | 1.166 (1.058, 1.284) | 0.002 | 0.005 | 1.119 (1.024, 1.223) | 0.013 | 0.059 | 1.040 (0.956, 1.131) | 0.367 | 1.919 |
| Concentration of medium VLDL particles | mol/l | 1.187 (1.078, 1.308) | <0.001 | 0.003 | 1.136 (1.037, 1.244) | 0.006 | 0.039 | 1.036 (0.952, 1.128) | 0.410 | 1.618 |
| Concentration of small VLDL particles | mol/l | 1.237 (1.122, 1.363) | <0.001 | <0.001 | 1.144 (1.043, 1.255) | 0.004 | 0.039 | 1.033 (0.949, 1.125) | 0.453 | 1.417 |
| Concentration of very small VLDL particles | mol/l | 1.247 (1.130, 1.376) | <0.001 | <0.001 | 1.121 (1.019, 1.233) | 0.019 | 0.074 | 1.004 (0.921, 1.095) | 0.922 | 1.012 |
| Concentration of IDL particles | mol/l | 1.173 (1.062, 1.296) | 0.002 | 0.005 | 1.093 (0.993, 1.203) | 0.070 | 0.161 | 0.982 (0.901, 1.071) | 0.684 | 1.077 |
| Concentration of large LDL particles | mol/l | 1.181 (1.069, 1.304) | 0.001 | 0.004 | 1.109 (1.008, 1.221) | 0.033 | 0.099 | 0.994 (0.911, 1.083) | 0.882 | 1.003 |
| Concentration of medium LDL particles | mol/l | 1.186 (1.074, 1.310) | 0.001 | 0.003 | 1.132 (1.029, 1.245) | 0.011 | 0.053 | 1.006 (0.923, 1.097) | 0.894 | 1.001 |
| Concentration of small LDL particles | mol/l | 1.197 (1.084, 1.322) | <0.001 | 0.002 | 1.152 (1.047, 1.269) | 0.004 | 0.039 | 1.010 (0.926, 1.101) | 0.828 | 1.018 |
| Concentration of very large HDL particles | mol/l | 0.940 (0.853, 1.035) | 0.204 | 0.271 | 0.974 (0.889, 1.067) | 0.572 | 0.691 | 1.001 (0.922, 1.088) | 0.977 | 1.018 |
| Concentration of large HDL particles | mol/l | 0.838 (0.758, 0.926) | 0.001 | 0.003 | 0.908 (0.826, 0.999) | 0.048 | 0.125 | 0.983 (0.900, 1.073) | 0.697 | 1.074 |
| Concentration of medium HDL particles | mol/l | 0.873 (0.791, 0.963) | 0.007 | 0.015 | 0.990 (0.898, 1.091) | 0.837 | 0.884 | 0.970 (0.889, 1.058) | 0.489 | 1.428 |
| Concentration of small HDL particles | mol/l | 1.022 (0.926, 1.127) | 0.669 | 0.738 | 1.071 (0.972, 1.180) | 0.165 | 0.291 | 1.003 (0.922, 1.092) | 0.940 | 1.007 |
| Total lipids in chylomicrons and extremely large VLDL | mmol/l | 1.099 (0.995, 1.213) | 0.063 | 0.104 | 1.079 (0.991, 1.176) | 0.081 | 0.177 | 1.014 (0.935, 1.100) | 0.739 | 1.046 |
| Total lipids in very large VLDL | mmol/l | 1.113 (1.011, 1.226) | 0.029 | 0.055 | 1.104 (1.010, 1.207) | 0.029 | 0.094 | 1.023 (0.941, 1.112) | 0.597 | 1.222 |
| Total lipids in large VLDL | mmol/l | 1.165 (1.057, 1.283) | 0.002 | 0.006 | 1.118 (1.022, 1.222) | 0.014 | 0.062 | 1.038 (0.955, 1.130) | 0.380 | 1.818 |
| Total lipids in medium VLDL | mmol/l | 1.188 (1.078, 1.308) | <0.001 | 0.003 | 1.135 (1.037, 1.243) | 0.006 | 0.039 | 1.036 (0.951, 1.128) | 0.418 | 1.543 |
| Total lipids in small VLDL | mmol/l | 1.241 (1.126, 1.368) | <0.001 | <0.001 | 1.146 (1.045, 1.258) | 0.004 | 0.037 | 1.033 (0.948, 1.125) | 0.456 | 1.407 |
| Total lipids in very small VLDL | mmol/l | 1.235 (1.119, 1.363) | <0.001 | <0.001 | 1.114 (1.012, 1.226) | 0.027 | 0.094 | 1.002 (0.919, 1.092) | 0.970 | 1.015 |
| Total lipids in IDL | mmol/l | 1.160 (1.050, 1.281) | 0.003 | 0.009 | 1.090 (0.991, 1.200) | 0.077 | 0.172 | 0.981 (0.899, 1.069) | 0.657 | 1.079 |
| Total lipids in large LDL | mmol/l | 1.171 (1.060, 1.293) | 0.002 | 0.005 | 1.106 (1.006, 1.217) | 0.038 | 0.107 | 0.992 (0.911, 1.082) | 0.864 | 0.997 |
| Total lipids in medium LDL | mmol/l | 1.180 (1.069, 1.303) | 0.001 | 0.004 | 1.130 (1.027, 1.243) | 0.012 | 0.056 | 1.005 (0.922, 1.096) | 0.905 | 1.008 |
| Total lipids in small LDL | mmol/l | 1.189 (1.077, 1.313) | 0.001 | 0.003 | 1.146 (1.042, 1.262) | 0.005 | 0.041 | 1.008 (0.925, 1.099) | 0.853 | 0.989 |
| Total lipids in very large HDL | mmol/l | 0.941 (0.855, 1.036) | 0.214 | 0.280 | 0.975 (0.890, 1.069) | 0.593 | 0.706 | 1.001 (0.922, 1.087) | 0.983 | 1.010 |
| Total lipids in large HDL | mmol/l | 0.836 (0.756, 0.924) | <0.001 | 0.003 | 0.908 (0.825, 0.999) | 0.047 | 0.122 | 0.982 (0.899, 1.072) | 0.685 | 1.070 |
| Total lipids in medium HDL | mmol/l | 0.868 (0.787, 0.958) | 0.005 | 0.012 | 0.989 (0.897, 1.090) | 0.824 | 0.878 | 0.971 (0.891, 1.059) | 0.512 | 1.354 |
| Total lipids in small HDL | mmol/l | 1.017 (0.922, 1.121) | 0.739 | 0.792 | 1.066 (0.968, 1.174) | 0.195 | 0.321 | 1.002 (0.921, 1.090) | 0.966 | 1.016 |
| Serum total cholesterol | mmol/l | 1.156 (1.048, 1.275) | 0.004 | 0.010 | 1.126 (1.022, 1.239) | 0.016 | 0.065 | 0.996 (0.913, 1.086) | 0.924 | 1.009 |
| Total cholesterol in VLDL | mmol/l | 1.227 (1.113, 1.352) | <0.001 | <0.001 | 1.158 (1.054, 1.271) | 0.002 | 0.028 | 1.025 (0.941, 1.117) | 0.570 | 1.309 |
| Total cholesterol in chylomicrons and extremely large VLDL | mmol/l | 1.108 (1.003, 1.225) | 0.043 | 0.076 | 1.074 (0.987, 1.170) | 0.099 | 0.204 | 1.014 (0.935, 1.100) | 0.736 | 1.062 |
| Total cholesterol in very large VLDL | mmol/l | 1.091 (0.990, 1.203) | 0.079 | 0.124 | 1.086 (0.994, 1.185) | 0.067 | 0.159 | 1.023 (0.941, 1.112) | 0.588 | 1.298 |
| Total cholesterol in large VLDL | mmol/l | 1.151 (1.043, 1.270) | 0.005 | 0.012 | 1.094 (1.002, 1.195) | 0.044 | 0.119 | 1.030 (0.947, 1.121) | 0.484 | 1.433 |
| Total cholesterol in medium VLDL | mmol/l | 1.183 (1.073, 1.303) | 0.001 | 0.003 | 1.121 (1.024, 1.226) | 0.013 | 0.058 | 1.027 (0.943, 1.118) | 0.546 | 1.322 |
| Total cholesterol in small VLDL | mmol/l | 1.247 (1.130, 1.375) | <0.001 | <0.001 | 1.146 (1.042, 1.260) | 0.005 | 0.040 | 1.030 (0.945, 1.123) | 0.495 | 1.427 |
| Total cholesterol in very small VLDL | mmol/l | 1.179 (1.067, 1.304) | 0.001 | 0.004 | 1.083 (0.984, 1.192) | 0.101 | 0.205 | 0.991 (0.910, 1.080) | 0.844 | 1.000 |
| Total cholesterol in IDL | mmol/l | 1.132 (1.025, 1.251) | 0.014 | 0.030 | 1.088 (0.989, 1.197) | 0.083 | 0.181 | 0.977 (0.896, 1.065) | 0.595 | 1.227 |
| Total cholesterol in LDL | mmol/l | 1.146 (1.036, 1.268) | 0.008 | 0.018 | 1.124 (1.018, 1.241) | 0.020 | 0.077 | 0.988 (0.910, 1.073) | 0.776 | 1.033 |

| | | | | | | | | | | |
|---|---|---|---|---|---|---|---|---|---|---|
| Total cholesterol in large LDL | mmol/l | 1.148 (1.038, 1.269) | 0.007 | 0.016 | 1.106 (1.005, 1.218) | 0.040 | 0.112 | 0.988 (0.909, 1.075) | 0.782 | 1.029 |
| Total cholesterol in medium LDL | mmol/l | 1.135 (1.029, 1.252) | 0.012 | 0.025 | 1.111 (1.011, 1.222) | 0.029 | 0.095 | 1.004 (0.921, 1.094) | 0.933 | 1.004 |
| Total cholesterol in small LDL | mmol/l | 1.130 (1.024, 1.246) | 0.015 | 0.030 | 1.113 (1.012, 1.223) | 0.028 | 0.097 | 1.005 (0.922, 1.095) | 0.913 | 1.012 |
| Total cholesterol in HDL | mmol/l | 0.863 (0.784, 0.951) | 0.003 | 0.008 | 0.958 (0.872, 1.054) | 0.381 | 0.523 | 0.981 (0.900, 1.069) | 0.666 | 1.078 |
| Total cholesterol in HDL2 | mmol/l | 0.841 (0.765, 0.926) | <0.001 | 0.002 | 0.951 (0.864, 1.045) | 0.296 | 0.436 | 0.972 (0.892, 1.060) | 0.521 | 1.332 |
| Total cholesterol in HDL3 | mmol/l | 1.099 (0.998, 1.211) | 0.056 | 0.093 | 1.065 (0.969, 1.170) | 0.193 | 0.318 | 1.060 (0.974, 1.153) | 0.175 | 2.629 |
| Total cholesterol in very large HDL | mmol/l | 0.993 (0.903, 1.092) | 0.885 | 0.913 | 1.016 (0.928, 1.113) | 0.731 | 0.807 | 1.013 (0.933, 1.100) | 0.762 | 1.032 |
| Total cholesterol in large HDL | mmol/l | 0.826 (0.748, 0.912) | <0.001 | 0.001 | 0.904 (0.822, 0.994) | 0.037 | 0.106 | 0.977 (0.895, 1.067) | 0.608 | 1.180 |
| Total cholesterol in medium HDL | mmol/l | 0.849 (0.772, 0.935) | 0.001 | 0.003 | 0.995 (0.904, 1.095) | 0.911 | 0.919 | 0.972 (0.893, 1.057) | 0.503 | 1.349 |
| Total cholesterol in small HDL | mmol/l | 1.039 (0.943, 1.145) | 0.436 | 0.525 | 1.050 (0.957, 1.151) | 0.301 | 0.440 | 1.011 (0.928, 1.101) | 0.804 | 1.022 |
| Remnant cholesterol (non-HDL, non-LDL -cholesterol) | mmol/l | 1.228 (1.113, 1.356) | <0.001 | <0.001 | 1.163 (1.056, 1.281) | 0.002 | 0.027 | 1.008 (0.924, 1.100) | 0.852 | 0.993 |
| Esterified cholesterol | mmol/l | 0.972 (0.897, 1.052) | 0.476 | 0.564 | 1.010 (0.853, 1.195) | 0.909 | 0.922 | 0.921 (0.682, 1.243) | 0.589 | 1.287 |
| Cholesterol esters in chylomicrons and extremely large VLDL | mmol/l | 1.110 (1.005, 1.227) | 0.040 | 0.072 | 1.074 (0.986, 1.170) | 0.104 | 0.207 | 1.012 (0.933, 1.097) | 0.777 | 1.028 |
| Cholesterol esters in very large VLDL | mmol/l | 1.082 (0.982, 1.193) | 0.111 | 0.165 | 1.074 (0.984, 1.172) | 0.109 | 0.214 | 1.024 (0.942, 1.113) | 0.579 | 1.290 |
| Cholesterol esters in large VLDL | mmol/l | 1.133 (1.022, 1.257) | 0.017 | 0.035 | 1.060 (0.975, 1.153) | 0.172 | 0.295 | 1.041 (0.957, 1.133) | 0.347 | 2.170 |
| Cholesterol esters in medium VLDL | mmol/l | 1.164 (1.056, 1.284) | 0.002 | 0.006 | 1.107 (1.011, 1.212) | 0.029 | 0.095 | 1.011 (0.927, 1.101) | 0.809 | 1.017 |
| Cholesterol esters in small VLDL | mmol/l | 1.211 (1.097, 1.338) | <0.001 | 0.001 | 1.131 (1.027, 1.246) | 0.012 | 0.057 | 1.022 (0.938, 1.114) | 0.620 | 1.134 |
| Cholesterol esters in very small VLDL | mmol/l | 1.166 (1.054, 1.289) | 0.003 | 0.008 | 1.082 (0.982, 1.192) | 0.109 | 0.216 | 0.988 (0.906, 1.077) | 0.784 | 1.019 |
| Cholesterol esters in IDL | mmol/l | 1.150 (1.041, 1.271) | 0.006 | 0.014 | 1.106 (1.005, 1.217) | 0.040 | 0.110 | 0.977 (0.896, 1.066) | 0.600 | 1.216 |
| Cholesterol esters in large LDL | mmol/l | 1.176 (1.051, 1.316) | 0.005 | 0.011 | 1.122 (1.014, 1.243) | 0.026 | 0.094 | 0.986 (0.912, 1.066) | 0.726 | 1.075 |
| Cholesterol esters in medium LDL | mmol/l | 1.082 (0.982, 1.193) | 0.111 | 0.165 | 1.091 (0.990, 1.201) | 0.077 | 0.171 | 0.999 (0.920, 1.085) | 0.981 | 1.018 |
| Cholesterol esters in small LDL | mmol/l | 1.069 (0.971, 1.176) | 0.175 | 0.237 | 1.084 (0.984, 1.195) | 0.101 | 0.206 | 1.000 (0.921, 1.085) | 0.996 | 0.996 |
| Cholesterol esters in very large HDL | mmol/l | 1.015 (0.923, 1.115) | 0.762 | 0.805 | 1.033 (0.943, 1.131) | 0.485 | 0.617 | 1.021 (0.941, 1.108) | 0.620 | 1.125 |
| Cholesterol esters in large HDL | mmol/l | 0.823 (0.745, 0.910) | <0.001 | 0.001 | 0.902 (0.820, 0.993) | 0.034 | 0.101 | 0.978 (0.895, 1.068) | 0.619 | 1.152 |
| Cholesterol esters in medium HDL | mmol/l | 0.848 (0.770, 0.933) | 0.001 | 0.003 | 0.999 (0.908, 1.099) | 0.984 | 0.984 | 0.970 (0.892, 1.055) | 0.478 | 1.434 |
| Cholesterol esters in small HDL | mmol/l | 1.021 (0.922, 1.131) | 0.688 | 0.755 | 1.043 (0.952, 1.142) | 0.367 | 0.512 | 0.989 (0.911, 1.074) | 0.794 | 1.020 |
| Free cholesterol | mmol/l | 1.110 (1.009, 1.222) | 0.033 | 0.060 | 1.152 (1.044, 1.271) | 0.005 | 0.041 | 0.978 (0.896, 1.067) | 0.610 | 1.163 |
| Free cholesterol in chylomicrons and extremely large VLDL | mmol/l | 1.087 (0.985, 1.199) | 0.097 | 0.149 | 1.081 (0.992, 1.177) | 0.075 | 0.169 | 1.018 (0.939, 1.105) | 0.661 | 1.078 |
| Free cholesterol in very large VLDL | mmol/l | 1.072 (0.973, 1.181) | 0.162 | 0.225 | 1.086 (0.996, 1.185) | 0.062 | 0.151 | 1.021 (0.940, 1.109) | 0.626 | 1.127 |
| Free cholesterol in large VLDL | mmol/l | 1.147 (1.039, 1.266) | 0.007 | 0.015 | 1.099 (1.008, 1.199) | 0.032 | 0.098 | 1.044 (0.960, 1.135) | 0.316 | 2.367 |
| Free cholesterol in medium VLDL | mmol/l | 1.189 (1.079, 1.310) | <0.001 | 0.003 | 1.127 (1.031, 1.232) | 0.009 | 0.051 | 1.040 (0.956, 1.132) | 0.364 | 2.050 |
| Free cholesterol in small VLDL | mmol/l | 1.250 (1.134, 1.377) | <0.001 | <0.001 | 1.139 (1.038, 1.250) | 0.006 | 0.040 | 1.034 (0.950, 1.126) | 0.443 | 1.467 |
| Free cholesterol in very small VLDL | mmol/l | 1.192 (1.076, 1.320) | 0.001 | 0.003 | 1.076 (0.981, 1.179) | 0.119 | 0.228 | 0.998 (0.919, 1.083) | 0.961 | 1.015 |
| Free cholesterol in IDL | mmol/l | 1.082 (0.980, 1.195) | 0.119 | 0.174 | 1.044 (0.950, 1.147) | 0.369 | 0.512 | 0.975 (0.897, 1.059) | 0.547 | 1.310 |
| Free cholesterol in large LDL | mmol/l | 1.112 (1.007, 1.228) | 0.037 | 0.066 | 1.065 (0.968, 1.171) | 0.195 | 0.318 | 0.980 (0.901, 1.067) | 0.643 | 1.113 |
| Free cholesterol in medium LDL | mmol/l | 1.178 (1.068, 1.301) | 0.001 | 0.004 | 1.121 (1.019, 1.234) | 0.019 | 0.075 | 0.999 (0.916, 1.090) | 0.988 | 1.002 |
| Free cholesterol in small LDL | mmol/l | 1.183 (1.072, 1.306) | 0.001 | 0.003 | 1.132 (1.029, 1.247) | 0.011 | 0.054 | 1.006 (0.923, 1.096) | 0.893 | 1.005 |
| Free cholesterol in very large HDL | mmol/l | 0.942 (0.857, 1.036) | 0.222 | 0.288 | 0.977 (0.892, 1.071) | 0.619 | 0.730 | 0.994 (0.915, 1.080) | 0.887 | 1.003 |
| Free cholesterol in large HDL | mmol/l | 0.846 (0.767, 0.934) | 0.001 | 0.004 | 0.916 (0.833, 1.007) | 0.069 | 0.162 | 0.976 (0.897, 1.062) | 0.572 | 1.300 |
| Free cholesterol in medium HDL | mmol/l | 0.864 (0.784, 0.952) | 0.003 | 0.008 | 0.978 (0.888, 1.077) | 0.646 | 0.746 | 0.978 (0.897, 1.067) | 0.619 | 1.171 |
| Free cholesterol in small HDL | mmol/l | 0.954 (0.865, 1.053) | 0.350 | 0.433 | 1.024 (0.929, 1.130) | 0.632 | 0.737 | 1.000 (0.919, 1.087) | 0.991 | 1.000 |
| Serum total triglycerides | mmol/l | 1.201 (1.091, 1.322) | <0.001 | 0.001 | 1.134 (1.035, 1.241) | 0.007 | 0.042 | 1.040 (0.955, 1.132) | 0.368 | 1.884 |
| Triglycerides in VLDL | mmol/l | 1.184 (1.075, 1.303) | 0.001 | 0.003 | 1.137 (1.038, 1.245) | 0.006 | 0.040 | 1.038 (0.954, 1.130) | 0.384 | 1.798 |
| Triglycerides in chylomicrons and extremely large VLDL | mmol/l | 1.089 (0.986, 1.204) | 0.093 | 0.144 | 1.068 (0.981, 1.161) | 0.129 | 0.243 | 1.013 (0.935, 1.098) | 0.748 | 1.038 |

| | | | | | | | | | | |
|---|---|---|---|---|---|---|---|---|---|---|
| Triglycerides in very large VLDL | mmol/l | 1.112 (1.010, 1.224) | 0.031 | 0.057 | 1.104 (1.010, 1.207) | 0.030 | 0.094 | 1.024 (0.942, 1.114) | 0.579 | 1.302 |
| Triglycerides in large VLDL | mmol/l | 1.167 (1.060, 1.285) | 0.002 | 0.005 | 1.123 (1.027, 1.228) | 0.011 | 0.054 | 1.044 (0.959, 1.135) | 0.321 | 2.329 |
| Triglycerides in medium VLDL | mmol/l | 1.186 (1.077, 1.306) | 0.001 | 0.003 | 1.138 (1.039, 1.246) | 0.005 | 0.040 | 1.040 (0.955, 1.132) | 0.364 | 2.101 |
| Triglycerides in small VLDL | mmol/l | 1.214 (1.102, 1.338) | <0.001 | 0.001 | 1.130 (1.031, 1.238) | 0.009 | 0.051 | 1.036 (0.952, 1.128) | 0.411 | 1.593 |
| Triglycerides in very small VLDL | mmol/l | 1.249 (1.133, 1.375) | <0.001 | <0.001 | 1.108 (1.011, 1.215) | 0.029 | 0.097 | 1.020 (0.937, 1.111) | 0.643 | 1.122 |
| Triglycerides in IDL | mmol/l | 1.272 (1.153, 1.404) | <0.001 | <0.001 | 1.071 (0.976, 1.176) | 0.148 | 0.275 | 1.014 (0.933, 1.103) | 0.737 | 1.056 |
| Triglycerides in LDL | mmol/l | 1.270 (1.150, 1.402) | <0.001 | <0.001 | 1.079 (0.983, 1.185) | 0.111 | 0.216 | 1.026 (0.943, 1.117) | 0.546 | 1.334 |
| Triglycerides in large LDL | mmol/l | 1.274 (1.152, 1.409) | <0.001 | <0.001 | 1.063 (0.967, 1.168) | 0.205 | 0.331 | 1.014 (0.933, 1.103) | 0.736 | 1.069 |
| Triglycerides in medium LDL | mmol/l | 1.255 (1.137, 1.385) | <0.001 | <0.001 | 1.096 (0.998, 1.203) | 0.055 | 0.141 | 1.028 (0.947, 1.115) | 0.513 | 1.343 |
| Triglycerides in small LDL | mmol/l | 1.266 (1.149, 1.396) | <0.001 | <0.001 | 1.155 (1.053, 1.268) | 0.002 | 0.026 | 1.037 (0.954, 1.126) | 0.393 | 1.732 |
| Triglycerides in HDL | mmol/l | 1.118 (1.012, 1.234) | 0.028 | 0.053 | 1.032 (0.941, 1.132) | 0.502 | 0.631 | 1.019 (0.934, 1.111) | 0.677 | 1.080 |
| Triglycerides in very large HDL | mmol/l | 1.125 (1.017, 1.246) | 0.023 | 0.045 | 1.043 (0.951, 1.143) | 0.370 | 0.511 | 1.045 (0.958, 1.139) | 0.324 | 2.277 |
| Triglycerides in large HDL | mmol/l | 1.014 (0.909, 1.132) | 0.802 | 0.840 | 0.935 (0.850, 1.028) | 0.165 | 0.292 | 1.020 (0.933, 1.116) | 0.657 | 1.087 |
| Triglycerides in medium HDL | mmol/l | 1.079 (0.981, 1.188) | 0.119 | 0.173 | 1.050 (0.958, 1.151) | 0.296 | 0.438 | 0.977 (0.897, 1.065) | 0.601 | 1.207 |
| Triglycerides in small HDL | mmol/l | 1.169 (1.062, 1.287) | 0.001 | 0.005 | 1.078 (0.985, 1.180) | 0.102 | 0.205 | 1.010 (0.929, 1.099) | 0.811 | 1.002 |
| Phospholipids in chylomicrons and extremely large VLDL | mmol/l | 1.089 (0.987, 1.201) | 0.090 | 0.141 | 1.082 (0.993, 1.180) | 0.072 | 0.164 | 1.014 (0.935, 1.099) | 0.738 | 1.051 |
| Phospholipids in very large VLDL | mmol/l | 1.082 (0.981, 1.192) | 0.113 | 0.167 | 1.084 (0.993, 1.183) | 0.072 | 0.163 | 1.021 (0.939, 1.109) | 0.630 | 1.125 |
| Phospholipids in large VLDL | mmol/l | 1.168 (1.059, 1.287) | 0.002 | 0.005 | 1.116 (1.022, 1.220) | 0.015 | 0.063 | 1.040 (0.956, 1.131) | 0.366 | 2.008 |
| Phospholipids in medium VLDL | mmol/l | 1.191 (1.082, 1.312) | <0.001 | 0.002 | 1.136 (1.038, 1.244) | 0.006 | 0.041 | 1.034 (0.949, 1.125) | 0.447 | 1.457 |
| Phospholipids in small VLDL | mmol/l | 1.227 (1.113, 1.351) | <0.001 | <0.001 | 1.131 (1.031, 1.241) | 0.009 | 0.052 | 1.020 (0.936, 1.111) | 0.655 | 1.092 |
| Phospholipids in very small VLDL | mmol/l | 1.181 (1.068, 1.306) | 0.001 | 0.004 | 1.095 (0.996, 1.204) | 0.059 | 0.147 | 0.988 (0.908, 1.075) | 0.783 | 1.025 |
| Phospholipids in IDL | mmol/l | 1.134 (1.027, 1.251) | 0.013 | 0.027 | 1.070 (0.974, 1.177) | 0.159 | 0.284 | 0.977 (0.896, 1.066) | 0.606 | 1.206 |
| Phospholipids in large LDL | mmol/l | 1.174 (1.063, 1.296) | 0.002 | 0.005 | 1.115 (1.013, 1.228) | 0.027 | 0.095 | 0.986 (0.904, 1.076) | 0.756 | 1.037 |
| Phospholipids in medium LDL | mmol/l | 1.223 (1.107, 1.350) | <0.001 | 0.001 | 1.164 (1.056, 1.284) | 0.002 | 0.027 | 0.995 (0.912, 1.086) | 0.913 | 1.007 |
| Phospholipids in small LDL | mmol/l | 1.216 (1.101, 1.342) | <0.001 | 0.001 | 1.174 (1.065, 1.296) | 0.001 | 0.020 | 1.000 (0.917, 1.090) | 0.994 | 0.999 |
| Phospholipids in very large HDL | mmol/l | 0.892 (0.810, 0.982) | 0.020 | 0.040 | 0.942 (0.860, 1.033) | 0.205 | 0.330 | 0.987 (0.908, 1.074) | 0.766 | 1.026 |
| Phospholipids in large HDL | mmol/l | 0.845 (0.765, 0.934) | 0.001 | 0.004 | 0.920 (0.837, 1.011) | 0.085 | 0.180 | 0.984 (0.902, 1.074) | 0.723 | 1.084 |
| Phospholipids in medium HDL | mmol/l | 0.885 (0.801, 0.978) | 0.016 | 0.034 | 0.980 (0.888, 1.080) | 0.678 | 0.770 | 0.980 (0.897, 1.070) | 0.651 | 1.109 |
| Phospholipids in small HDL | mmol/l | 0.950 (0.861, 1.047) | 0.299 | 0.378 | 1.030 (0.935, 1.134) | 0.549 | 0.675 | 0.989 (0.910, 1.076) | 0.801 | 1.024 |
| Apolipoprotein A-I | g/l | 0.965 (0.875, 1.064) | 0.477 | 0.562 | 1.037 (0.942, 1.142) | 0.455 | 0.585 | 1.011 (0.928, 1.101) | 0.810 | 1.013 |
| Apolipoprotein B | g/l | 1.260 (1.142, 1.390) | <0.001 | <0.001 | 1.200 (1.089, 1.323) | <0.001 | 0.009 | 1.021 (0.935, 1.115) | 0.649 | 1.114 |
| Ratio of apolipoprotein B to apolipoprotein A-I | | 1.274 (1.154, 1.406) | <0.001 | <0.001 | 1.176 (1.067, 1.296) | 0.001 | 0.019 | 1.016 (0.930, 1.110) | 0.723 | 1.091 |
| Total fatty acids | mmol/l | 1.151 (1.046, 1.266) | 0.004 | 0.010 | 1.221 (1.107, 1.347) | <0.001 | 0.003 | 1.022 (0.937, 1.115) | 0.620 | 1.143 |
| Saturated fatty acids | mmol/l | 1.132 (1.030, 1.244) | 0.010 | 0.021 | 1.191 (1.081, 1.311) | <0.001 | 0.011 | 1.011 (0.927, 1.102) | 0.806 | 1.019 |
| Monounsaturated fatty acids; 16:1, 18:1 | mmol/l | 1.178 (1.071, 1.296) | 0.001 | 0.003 | 1.185 (1.077, 1.303) | <0.001 | 0.011 | 1.030 (0.945, 1.122) | 0.500 | 1.407 |
| Polyunsaturated fatty acids | mmol/l | 1.119 (1.015, 1.233) | 0.024 | 0.047 | 1.273 (1.148, 1.412) | <0.001 | <0.001 | 1.024 (0.938, 1.119) | 0.593 | 1.270 |
| Omega-3 fatty acids | mmol/l | 1.170 (1.060, 1.292) | 0.002 | 0.005 | 1.193 (1.078, 1.321) | 0.001 | 0.013 | 0.970 (0.887, 1.060) | 0.500 | 1.390 |
| 22:6, docosahexaenoic acid | mmol/l | 1.213 (1.102, 1.335) | <0.001 | 0.001 | 1.160 (1.050, 1.282) | 0.004 | 0.039 | 0.987 (0.904, 1.077) | 0.761 | 1.037 |
| Omega-6 fatty acids | mmol/l | 1.104 (1.002, 1.216) | 0.045 | 0.078 | 1.277 (1.151, 1.416) | <0.001 | <0.001 | 1.034 (0.948, 1.129) | 0.448 | 1.440 |
| 18:2, linoleic acid | mmol/l | 0.942 (0.855, 1.037) | 0.222 | 0.287 | 1.147 (0.954, 1.379) | 0.144 | 0.269 | 0.973 (0.891, 1.063) | 0.543 | 1.343 |
| Ratio of saturated fatty acids to total fatty acids | % | 0.911 (0.829, 1.002) | 0.054 | 0.092 | 0.885 (0.805, 0.972) | 0.010 | 0.054 | 0.975 (0.887, 1.071) | 0.593 | 1.258 |
| Ratio of monounsaturated fatty acids to total fatty acids | % | 1.152 (1.046, 1.269) | 0.004 | 0.010 | 1.006 (0.916, 1.105) | 0.899 | 0.915 | 1.047 (0.960, 1.142) | 0.295 | 2.553 |
| Ratio of polyunsaturated fatty acids to total fatty acids | % | 0.932 (0.846, 1.027) | 0.156 | 0.218 | 1.066 (0.971, 1.172) | 0.180 | 0.304 | 1.022 (0.934, 1.118) | 0.637 | 1.119 |
| Ratio of omega-3 fatty acids to total fatty acids | % | 1.072 (0.972, 1.182) | 0.166 | 0.229 | 1.018 (0.921, 1.125) | 0.725 | 0.807 | 0.929 (0.849, 1.016) | 0.107 | 2.005 |

| | | | | | | | | | | |
|---|---|---|---|---|---|---|---|---|---|---|
| Ratio of 22:6 docosahexaenoic acid to total fatty acids | % | 1.064 (0.973, 1.165) | 0.173 | 0.236 | 0.937 (0.856, 1.025) | 0.154 | 0.278 | 0.971 (0.892, 1.058) | 0.503 | 1.364 |
| Ratio of omega-6 fatty acids to total fatty acids | % | 0.922 (0.839, 1.014) | 0.093 | 0.143 | 1.056 (0.965, 1.157) | 0.237 | 0.370 | 1.050 (0.961, 1.147) | 0.284 | 2.778 |
| Ratio of 18:2 linoleic acid to total fatty acids | % | 0.933 (0.853, 1.020) | 0.125 | 0.181 | 0.994 (0.908, 1.087) | 0.888 | 0.908 | 1.010 (0.921, 1.107) | 0.837 | 1.002 |
| Alanine | mmol/l | 1.006 (0.914, 1.108) | 0.902 | 0.927 | 1.022 (0.936, 1.116) | 0.625 | 0.732 | 0.968 (0.891, 1.051) | 0.439 | 1.495 |
| Glutamine | mmol/l | 0.899 (0.815, 0.993) | 0.036 | 0.066 | 0.970 (0.882, 1.067) | 0.532 | 0.661 | 0.966 (0.888, 1.050) | 0.416 | 1.561 |
| Histidine | mmol/l | 0.977 (0.889, 1.075) | 0.638 | 0.711 | 1.035 (0.948, 1.130) | 0.443 | 0.580 | 1.034 (0.954, 1.120) | 0.422 | 1.484 |
| Isoleucine | mmol/l | 1.018 (0.922, 1.123) | 0.728 | 0.791 | 1.031 (0.941, 1.130) | 0.510 | 0.637 | 0.955 (0.876, 1.040) | 0.289 | 2.603 |
| Leucine | mmol/l | 1.026 (0.927, 1.137) | 0.617 | 0.697 | 1.028 (0.934, 1.131) | 0.576 | 0.690 | 0.924 (0.847, 1.008) | 0.075 | 1.677 |
| Phenylalanine | mmol/l | 1.068 (0.965, 1.182) | 0.206 | 0.270 | 0.935 (0.849, 1.030) | 0.171 | 0.297 | 1.089 (1.001, 1.185) | 0.046 | 1.490 |
| Tyrosine | mmol/l | 0.976 (0.884, 1.078) | 0.637 | 0.713 | 0.986 (0.899, 1.082) | 0.769 | 0.836 | 1.046 (0.963, 1.136) | 0.284 | 2.665 |
| Valine | mmol/l | 0.997 (0.901, 1.102) | 0.950 | 0.967 | 1.028 (0.936, 1.129) | 0.560 | 0.685 | 0.908 (0.834, 0.987) | 0.024 | 1.071 |
| Citrate | mmol/l | 0.959 (0.870, 1.057) | 0.401 | 0.488 | 0.914 (0.832, 1.004) | 0.061 | 0.149 | 0.960 (0.884, 1.044) | 0.341 | 2.195 |
| Glucose | mmol/l | 1.321 (1.196, 1.459) | <0.001 | <0.001 | 1.180 (1.068, 1.305) | 0.001 | 0.019 | 1.309 (1.191, 1.439) | <0.001 | <0.001 |
| Lactate | mmol/l | 1.087 (0.983, 1.202) | 0.104 | 0.157 | 1.011 (0.922, 1.110) | 0.809 | 0.867 | 1.014 (0.931, 1.104) | 0.754 | 1.041 |
| Glycoprotein acetyls, mainly a1-acid glycoprotein | mmol/l | 1.334 (1.205, 1.477) | <0.001 | <0.001 | 1.274 (1.157, 1.403) | <0.001 | <0.001 | 1.157 (1.057, 1.266) | 0.002 | 0.114 |
| Acetoacetate | mmol/l | 0.936 (0.846, 1.035) | 0.196 | 0.262 | 1.234 (1.117, 1.363) | <0.001 | 0.002 | 1.051 (0.961, 1.149) | 0.277 | 2.836 |
| Acetate | mmol/l | 1.047 (0.945, 1.161) | 0.381 | 0.468 | 0.956 (0.859, 1.064) | 0.414 | 0.547 | 1.033 (0.949, 1.124) | 0.451 | 1.430 |
| 3-hydroxybutyrate | mmol/l | 1.214 (1.085, 1.358) | 0.001 | 0.003 | 1.134 (1.024, 1.256) | 0.015 | 0.064 | 1.093 (0.994, 1.202) | 0.066 | 1.640 |
| Cholesterol esters to total lipids ratio in IDL | % | 1.030 (0.932, 1.137) | 0.564 | 0.647 | 1.133 (1.031, 1.245) | 0.009 | 0.053 | 0.961 (0.886, 1.042) | 0.338 | 2.233 |
| Cholesterol esters to total lipids ratio in chylomicrons and extremely large VLDL | % | 1.103 (0.998, 1.219) | 0.054 | 0.091 | 1.043 (0.958, 1.136) | 0.332 | 0.473 | 1.010 (0.932, 1.094) | 0.810 | 1.007 |
| Cholesterol esters to total lipids ratio in large HDL | % | 0.808 (0.734, 0.890) | <0.001 | <0.001 | 0.901 (0.820, 0.991) | 0.032 | 0.099 | 0.963 (0.883, 1.050) | 0.391 | 1.757 |
| Cholesterol esters to total lipids ratio in large LDL | % | 1.152 (0.972, 1.365) | 0.104 | 0.157 | 1.123 (0.984, 1.281) | 0.085 | 0.179 | 0.984 (0.919, 1.054) | 0.652 | 1.104 |
| Cholesterol esters to total lipids ratio in large VLDL | % | 1.035 (0.936, 1.143) | 0.505 | 0.589 | 0.993 (0.917, 1.076) | 0.868 | 0.892 | 1.034 (0.951, 1.124) | 0.439 | 1.474 |
| Cholesterol esters to total lipids ratio in medium HDL | % | 0.851 (0.771, 0.940) | 0.001 | 0.005 | 1.021 (0.932, 1.119) | 0.649 | 0.745 | 0.979 (0.904, 1.060) | 0.594 | 1.238 |
| Cholesterol esters to total lipids ratio in medium LDL | % | 1.017 (0.926, 1.117) | 0.719 | 0.785 | 1.054 (0.957, 1.161) | 0.290 | 0.432 | 0.996 (0.920, 1.079) | 0.930 | 1.006 |
| Cholesterol esters to total lipids ratio in medium VLDL | % | 0.957 (0.865, 1.059) | 0.400 | 0.489 | 0.941 (0.855, 1.037) | 0.220 | 0.349 | 0.942 (0.863, 1.028) | 0.181 | 2.547 |
| Cholesterol esters to total lipids ratio in small HDL | % | 1.017 (0.918, 1.127) | 0.745 | 0.794 | 1.027 (0.937, 1.126) | 0.572 | 0.689 | 0.987 (0.909, 1.072) | 0.764 | 1.030 |
| Cholesterol esters to total lipids ratio in small LDL | % | 1.012 (0.922, 1.111) | 0.798 | 0.839 | 1.043 (0.948, 1.149) | 0.387 | 0.525 | 0.997 (0.921, 1.080) | 0.947 | 1.005 |
| Cholesterol esters to total lipids ratio in small VLDL | % | 0.998 (0.904, 1.100) | 0.961 | 0.965 | 0.991 (0.902, 1.088) | 0.844 | 0.884 | 0.993 (0.913, 1.080) | 0.874 | 0.998 |
| Cholesterol esters to total lipids ratio in very large HDL | % | 1.249 (1.131, 1.380) | <0.001 | <0.001 | 1.190 (1.080, 1.312) | <0.001 | 0.011 | 1.064 (0.974, 1.161) | 0.168 | 2.705 |
| Cholesterol esters to total lipids ratio in very large VLDL | % | 0.990 (0.900, 1.088) | 0.830 | 0.865 | 1.001 (0.919, 1.091) | 0.975 | 0.979 | 1.020 (0.940, 1.107) | 0.631 | 1.119 |
| Cholesterol esters to total lipids ratio in very small VLDL | % | 0.898 (0.815, 0.990) | 0.031 | 0.057 | 0.948 (0.860, 1.045) | 0.282 | 0.426 | 0.960 (0.881, 1.047) | 0.359 | 2.182 |
| Free cholesterol to total lipids ratio in IDL | % | 0.949 (0.857, 1.051) | 0.312 | 0.390 | 0.974 (0.884, 1.074) | 0.600 | 0.710 | 0.966 (0.894, 1.045) | 0.393 | 1.702 |
| Free cholesterol to total lipids ratio in chylomicrons and extremely large VLDL | % | 1.069 (0.968, 1.181) | 0.186 | 0.250 | 1.058 (0.973, 1.150) | 0.187 | 0.313 | 1.024 (0.945, 1.109) | 0.567 | 1.316 |
| Free cholesterol to total lipids ratio in large HDL | % | 0.901 (0.816, 0.995) | 0.039 | 0.069 | 0.955 (0.869, 1.049) | 0.337 | 0.477 | 0.974 (0.903, 1.050) | 0.496 | 1.411 |
| Free cholesterol to total lipids ratio in large LDL | % | 0.846 (0.765, 0.935) | 0.001 | 0.004 | 0.876 (0.793, 0.969) | 0.010 | 0.053 | 0.953 (0.877, 1.034) | 0.245 | 2.906 |
| Free cholesterol to total lipids ratio in large VLDL | % | 1.100 (0.993, 1.218) | 0.067 | 0.109 | 1.063 (0.978, 1.154) | 0.151 | 0.277 | 1.048 (0.965, 1.139) | 0.262 | 2.808 |
| Free cholesterol to total lipids ratio in medium HDL | % | 0.876 (0.796, 0.963) | 0.006 | 0.015 | 0.956 (0.871, 1.049) | 0.344 | 0.484 | 0.991 (0.910, 1.079) | 0.840 | 1.000 |
| Free cholesterol to total lipids ratio in medium LDL | % | 0.876 (0.794, 0.967) | 0.009 | 0.019 | 0.899 (0.818, 0.989) | 0.028 | 0.095 | 0.991 (0.910, 1.080) | 0.834 | 1.015 |
| Free cholesterol to total lipids ratio in medium VLDL | % | 1.179 (1.063, 1.308) | 0.002 | 0.005 | 1.077 (0.989, 1.173) | 0.088 | 0.183 | 1.057 (0.972, 1.149) | 0.195 | 2.587 |
| Free cholesterol to total lipids ratio in small HDL | % | 0.883 (0.799, 0.976) | 0.015 | 0.030 | 0.941 (0.852, 1.040) | 0.235 | 0.369 | 0.991 (0.908, 1.083) | 0.850 | 0.996 |
| Free cholesterol to total lipids ratio in small LDL | % | 0.888 (0.804, 0.980) | 0.018 | 0.036 | 0.889 (0.808, 0.979) | 0.017 | 0.068 | 0.997 (0.916, 1.085) | 0.942 | 1.005 |
| Free cholesterol to total lipids ratio in small VLDL | % | 1.035 (0.937, 1.143) | 0.499 | 0.585 | 0.949 (0.865, 1.041) | 0.264 | 0.403 | 1.007 (0.927, 1.094) | 0.865 | 0.993 |
| Free cholesterol to total lipids ratio in very large HDL | % | 0.975 (0.891, 1.068) | 0.592 | 0.672 | 0.991 (0.903, 1.088) | 0.854 | 0.885 | 0.964 (0.884, 1.051) | 0.403 | 1.649 |

| | | | | | | | | | | |
|---|---|---|---|---|---|---|---|---|---|---|
| Free cholesterol to total lipids ratio in very large VLDL | % | 1.029 (0.935, 1.133) | 0.561 | 0.647 | 1.064 (0.977, 1.160) | 0.154 | 0.279 | 1.017 (0.938, 1.104) | 0.682 | 1.081 |
| Free cholesterol to total lipids ratio in very small VLDL | % | 1.006 (0.903, 1.120) | 0.917 | 0.938 | 0.986 (0.901, 1.080) | 0.769 | 0.839 | 0.986 (0.915, 1.063) | 0.717 | 1.089 |
| Phospholipids to total lipds ratio in IDL | % | 0.845 (0.765, 0.935) | 0.001 | 0.004 | 0.906 (0.822, 0.998) | 0.046 | 0.122 | 0.952 (0.876, 1.033) | 0.239 | 2.985 |
| Phospholipids to total lipds ratio in chylomicrons and extremely large VLDL | % | 1.071 (0.971, 1.182) | 0.169 | 0.232 | 1.060 (0.974, 1.153) | 0.175 | 0.298 | 1.014 (0.937, 1.097) | 0.728 | 1.064 |
| Phospholipids to total lipds ratio in large HDL | % | 1.202 (1.085, 1.331) | <0.001 | 0.003 | 1.140 (1.036, 1.255) | 0.007 | 0.044 | 1.028 (0.945, 1.118) | 0.526 | 1.329 |
| Phospholipids to total lipds ratio in large LDL | % | 0.883 (0.802, 0.972) | 0.011 | 0.024 | 0.935 (0.852, 1.025) | 0.151 | 0.279 | 0.964 (0.886, 1.049) | 0.395 | 1.678 |
| Phospholipids to total lipds ratio in large VLDL | % | 1.158 (1.043, 1.285) | 0.006 | 0.014 | 1.083 (0.995, 1.179) | 0.065 | 0.156 | 1.039 (0.956, 1.129) | 0.367 | 1.964 |
| Phospholipids to total lipds ratio in medium HDL | % | 1.190 (1.076, 1.315) | 0.001 | 0.003 | 0.950 (0.861, 1.048) | 0.306 | 0.442 | 1.048 (0.967, 1.137) | 0.255 | 2.868 |
| Phospholipids to total lipds ratio in medium LDL | % | 0.944 (0.858, 1.039) | 0.240 | 0.308 | 0.962 (0.878, 1.054) | 0.403 | 0.540 | 0.984 (0.904, 1.072) | 0.713 | 1.091 |
| Phospholipids to total lipds ratio in medium VLDL | % | 1.000 (0.909, 1.099) | 0.992 | 0.992 | 0.966 (0.883, 1.056) | 0.446 | 0.579 | 0.937 (0.862, 1.019) | 0.127 | 2.198 |
| Phospholipids to total lipds ratio in small HDL | % | 0.895 (0.812, 0.986) | 0.025 | 0.049 | 0.968 (0.881, 1.063) | 0.492 | 0.621 | 0.974 (0.893, 1.062) | 0.547 | 1.283 |
| Phospholipids to total lipds ratio in small LDL | % | 0.932 (0.847, 1.025) | 0.149 | 0.212 | 0.947 (0.863, 1.039) | 0.246 | 0.382 | 0.991 (0.910, 1.079) | 0.831 | 1.016 |
| Phospholipids to total lipds ratio in small VLDL | % | 0.824 (0.746, 0.911) | <0.001 | 0.001 | 0.844 (0.768, 0.926) | <0.001 | 0.012 | 0.909 (0.834, 0.991) | 0.030 | 1.137 |
| Phospholipids to total lipds ratio in very large HDL | % | 0.832 (0.756, 0.917) | <0.001 | 0.001 | 0.893 (0.811, 0.982) | 0.020 | 0.076 | 0.964 (0.884, 1.051) | 0.400 | 1.667 |
| Phospholipids to total lipds ratio in very large VLDL | % | 1.024 (0.930, 1.128) | 0.626 | 0.704 | 1.045 (0.960, 1.138) | 0.311 | 0.446 | 1.018 (0.938, 1.105) | 0.667 | 1.071 |
| Phospholipids to total lipds ratio in very small VLDL | % | 0.997 (0.902, 1.102) | 0.955 | 0.968 | 1.018 (0.928, 1.117) | 0.699 | 0.782 | 0.969 (0.896, 1.047) | 0.423 | 1.464 |
| Total cholesterol to total lipids ratio in IDL | % | 0.969 (0.877, 1.071) | 0.536 | 0.622 | 1.071 (0.971, 1.181) | 0.169 | 0.294 | 0.959 (0.885, 1.039) | 0.302 | 2.343 |
| Total cholesterol to total lipids ratio in chylomicrons and extremely large VLDL | % | 1.108 (1.001, 1.225) | 0.047 | 0.082 | 1.045 (0.961, 1.137) | 0.305 | 0.442 | 1.014 (0.937, 1.099) | 0.724 | 1.079 |
| Total cholesterol to total lipids ratio in large HDL | % | 0.816 (0.742, 0.898) | <0.001 | <0.001 | 0.903 (0.821, 0.993) | 0.035 | 0.102 | 0.961 (0.883, 1.047) | 0.364 | 2.155 |
| Total cholesterol to total lipids ratio in large LDL | % | 1.041 (0.937, 1.156) | 0.452 | 0.542 | 1.084 (0.977, 1.202) | 0.127 | 0.241 | 0.980 (0.908, 1.057) | 0.594 | 1.250 |
| Total cholesterol to total lipids ratio in large VLDL | % | 1.062 (0.960, 1.176) | 0.243 | 0.311 | 1.012 (0.933, 1.098) | 0.778 | 0.841 | 1.019 (0.938, 1.108) | 0.655 | 1.099 |
| Total cholesterol to total lipids ratio in medium HDL | % | 0.844 (0.767, 0.929) | 0.001 | 0.003 | 1.009 (0.920, 1.106) | 0.850 | 0.886 | 0.979 (0.904, 1.061) | 0.610 | 1.173 |
| Total cholesterol to total lipids ratio in medium LDL | % | 1.017 (0.924, 1.119) | 0.732 | 0.788 | 1.054 (0.958, 1.159) | 0.280 | 0.426 | 1.001 (0.921, 1.088) | 0.985 | 1.007 |
| Total cholesterol to total lipids ratio in medium VLDL | % | 1.023 (0.926, 1.131) | 0.653 | 0.724 | 0.984 (0.898, 1.078) | 0.730 | 0.809 | 0.969 (0.890, 1.055) | 0.467 | 1.421 |
| Total cholesterol to total lipids ratio in small HDL | % | 1.036 (0.940, 1.141) | 0.475 | 0.565 | 1.019 (0.929, 1.118) | 0.695 | 0.782 | 1.009 (0.926, 1.099) | 0.836 | 1.006 |
| Total cholesterol to total lipids ratio in small LDL | % | 1.016 (0.924, 1.117) | 0.746 | 0.792 | 1.039 (0.945, 1.141) | 0.432 | 0.569 | 1.001 (0.920, 1.088) | 0.988 | 1.006 |
| Total cholesterol to total lipids ratio in small VLDL | % | 1.002 (0.909, 1.105) | 0.960 | 0.969 | 0.984 (0.898, 1.079) | 0.738 | 0.810 | 0.996 (0.915, 1.084) | 0.927 | 1.007 |
| Total cholesterol to total lipids ratio in very large HDL | % | 1.211 (1.098, 1.337) | <0.001 | 0.001 | 1.166 (1.059, 1.285) | 0.002 | 0.026 | 1.047 (0.960, 1.143) | 0.301 | 2.415 |
| Total cholesterol to total lipids ratio in very large VLDL | % | 1.010 (0.917, 1.111) | 0.846 | 0.877 | 1.025 (0.941, 1.117) | 0.566 | 0.688 | 1.022 (0.941, 1.109) | 0.608 | 1.189 |
| Total cholesterol to total lipids ratio in very small VLDL | % | 0.911 (0.825, 1.006) | 0.064 | 0.105 | 0.946 (0.858, 1.042) | 0.260 | 0.401 | 0.964 (0.886, 1.048) | 0.390 | 1.793 |
| Triglycerides to total lipids ratio in IDL | % | 1.099 (0.997, 1.211) | 0.057 | 0.094 | 0.990 (0.902, 1.088) | 0.839 | 0.882 | 1.023 (0.939, 1.114) | 0.606 | 1.197 |
| Triglycerides to total lipids ratio in chylomicrons and extremely large VLDL | % | 1.056 (0.957, 1.165) | 0.282 | 0.358 | 1.035 (0.954, 1.124) | 0.406 | 0.541 | 1.011 (0.934, 1.094) | 0.789 | 1.020 |
| Triglycerides to total lipids ratio in large HDL | % | 1.186 (1.068, 1.316) | 0.001 | 0.005 | 0.992 (0.906, 1.087) | 0.867 | 0.895 | 1.040 (0.953, 1.135) | 0.379 | 1.855 |
| Triglycerides to total lipids ratio in large LDL | % | 1.074 (0.973, 1.185) | 0.156 | 0.219 | 0.958 (0.871, 1.054) | 0.382 | 0.521 | 1.014 (0.931, 1.105) | 0.746 | 1.042 |
| Triglycerides to total lipids ratio in large VLDL | % | 0.973 (0.881, 1.073) | 0.581 | 0.664 | 0.990 (0.908, 1.080) | 0.828 | 0.879 | 1.088 (1.001, 1.182) | 0.047 | 1.310 |
| Triglycerides to total lipids ratio in medium HDL | % | 1.159 (1.054, 1.274) | 0.002 | 0.006 | 1.051 (0.959, 1.151) | 0.287 | 0.431 | 0.991 (0.910, 1.079) | 0.836 | 1.011 |
| Triglycerides to total lipids ratio in medium LDL | % | 1.066 (0.967, 1.175) | 0.201 | 0.267 | 0.972 (0.884, 1.068) | 0.549 | 0.679 | 1.027 (0.943, 1.118) | 0.543 | 1.358 |
| Triglycerides to total lipids ratio in medium VLDL | % | 0.982 (0.888, 1.086) | 0.730 | 0.789 | 1.012 (0.922, 1.112) | 0.800 | 0.861 | 1.037 (0.952, 1.129) | 0.405 | 1.628 |
| Triglycerides to total lipids ratio in small HDL | % | 1.165 (1.059, 1.282) | 0.002 | 0.005 | 1.059 (0.967, 1.159) | 0.214 | 0.342 | 1.008 (0.927, 1.097) | 0.850 | 1.001 |
| Triglycerides to total lipids ratio in small LDL | % | 1.098 (0.999, 1.207) | 0.053 | 0.091 | 1.034 (0.944, 1.132) | 0.472 | 0.603 | 1.036 (0.951, 1.127) | 0.419 | 1.520 |
| Triglycerides to total lipids ratio in small VLDL | % | 1.053 (0.955, 1.161) | 0.302 | 0.380 | 1.040 (0.950, 1.139) | 0.397 | 0.535 | 1.030 (0.945, 1.121) | 0.502 | 1.378 |
| Triglycerides to total lipids ratio in very large HDL | % | 1.168 (1.061, 1.287) | 0.002 | 0.005 | 1.061 (0.971, 1.160) | 0.192 | 0.319 | 1.047 (0.961, 1.140) | 0.296 | 2.469 |
| Triglycerides to total lipids ratio in very large VLDL | % | 0.924 (0.840, 1.017) | 0.105 | 0.158 | 0.967 (0.888, 1.054) | 0.446 | 0.576 | 1.034 (0.954, 1.120) | 0.421 | 1.505 |
| Triglycerides to total lipids ratio in very small VLDL | % | 1.074 (0.974, 1.183) | 0.151 | 0.214 | 1.022 (0.933, 1.119) | 0.645 | 0.748 | 1.015 (0.932, 1.106) | 0.727 | 1.070 |

| | | | | | | | | | |
|---|---|---|---|---|---|---|---|---|---|
| Total cholines | mmol/l | 1.074 (0.977, 1.181) | 0.138 | 0.197 | 1.152 (1.044, 1.271) | 0.005 | 0.041 | 1.026 (0.943, 1.116) | 0.547 | 1.296 |
| Phosphatidylcholine and other cholines | mmol/l | 1.048 (0.954, 1.151) | 0.327 | 0.406 | 1.118 (1.015, 1.232) | 0.024 | 0.090 | 1.001 (0.920, 1.089) | 0.982 | 1.014 |
| Total phosphoglycerides | mmol/l | 1.090 (0.991, 1.199) | 0.075 | 0.120 | 1.153 (1.047, 1.270) | 0.004 | 0.038 | 1.028 (0.945, 1.119) | 0.515 | 1.333 |
| Ratio of triglycerides to phosphoglycerides | | 1.149 (1.045, 1.264) | 0.004 | 0.011 | 1.126 (1.029, 1.232) | 0.010 | 0.051 | 1.040 (0.955, 1.132) | 0.369 | 1.847 |
| Albumin | signal area | 0.867 (0.785, 0.958) | 0.005 | 0.012 | 0.909 (0.828, 0.998) | 0.045 | 0.121 | 0.868 (0.797, 0.947) | 0.001 | 0.155 |
| Creatinine | mmol/l | 1.232 (1.083, 1.402) | 0.002 | 0.005 | 0.973 (0.852, 1.110) | 0.680 | 0.769 | 1.168 (1.056, 1.293) | 0.003 | 0.148 |
| Estimated degree of unsaturation | | 0.899 (0.819, 0.987) | 0.026 | 0.049 | 1.021 (0.931, 1.119) | 0.663 | 0.758 | 0.920 (0.839, 1.009) | 0.078 | 1.602 |
| Sphingomyelins | mmol/l | 1.041 (0.947, 1.145) | 0.407 | 0.492 | 1.100 (0.997, 1.213) | 0.057 | 0.144 | 0.978 (0.895, 1.068) | 0.619 | 1.161 |

Models were adjusted for age, sex, region, education, household income, occupation, marital status, tea drinking habit, smoking status, alcohol intake, physical activity, self-rated health, fasting time and frequency of 12 food groups.
